# Using Fisher Information Matrix to predict uncertainty in covariate effects and power to detect their relevance in Non-Linear Mixed Effect Models in pharmacometrics

**DOI:** 10.1101/2024.10.16.24314758

**Authors:** Lucie Fayette, Karl Brendel, France Mentré

## Abstract

This work focuses on design of experiments for Pharmacokinetic (PK) and Pharmacodynamic (PD) studies. Non-Linear Mixed effects Models (NLMEM) modeling allows the identification and quantification of covariates that explain inter-individual variability (IIV). The Fisher Information Matrix (FIM), computed by linearization, has already been used to predict uncertainty on covariate parameters and the power of a test to detect statistical significance. A covariate effect on a parameter is deemed statistically significant if it is different from 0 according to a Wald comparison test and clinically relevant if the ratio of change it causes in the parameter is relevant according to two one-sided tests (TOST) as in bioequivalence studies. FIM calculation was extended by computing its expectation on the joint distribution of the covariates, discrete and continuous. Three methods were proposed: using a provided sample of covariate vectors, simulating covariate vectors, based on provided independent distributions or on estimated copulas. Thereafter, CI of ratios, power of tests and number of subjects needed to achieve desired confidence were derived. Methods were implemented in a working version of the R package *PFIM*. A simulation study was conducted under various scenarios, including different sample sizes, sampling points, and IIV. Overall, uncertainty on covariate effects and power of tests were accurately predicted. The method was applied to a population PK model of the drug cabozantinib including 27 covariate relationships. Despite numerous relationships, limited representation of certain covariates, PFIM correctly predicted uncertainty, and is therefore suitable for rapidly computing number of subjects needed to achieve given powers.

## 1 Introduction

One of the main aims of pharmacokinetic (PK) and pharmacodynamic (PD) analysis is to identify and quantify the covariates that attempt to explain inter-individual variability (IIV). In this context Non-Linear Mixed effects Models (NLMEM) are an appropriate tool as population modeling allows to quantitatively describe relationships between covariates and parameters, and consequently to explain some of the IIV. [1] The list of covariates of interest can be represented on a forest plot where the 90% confidence interval (CI) of the ratio of change in the value of the parameter is expressed for given values, or categories, of the covariate, and relative to a reference value [2]. These plots are in line with the FDA Guidance for Industry Population Pharmacokinetics (2022) [3] and are essential to support clinicians’ decisions on drug dosing [4]. Therefore, regarding the pharmaceutical development of a new drug, it is essential to be able to detect covariates that may explain some of the PKPD variability, and to distinguish between a statistically significant and a clinically relevant effect. A covariate effect on a parameter is considered statistically significant if it is different from 0 according to a Wald test at level 95%, clinically relevant if the ratio of change it causes in the parameter is relevant according to a Two One-Sided Tests Procedure (TOST) at level 95%. The latter is equivalent to showing that the 90% CI of the ratio is outside a predetermined interval (often [0.80; 1.25]) [5]. The ratio is said irrelevant if this CI is included in the range. In the case of partial overlap, there is insufficient data to determine clinical relevance.

For a given model and parameters, the Standard Error (SE) of the covariate parameters and the width of CI of the ratio are influenced by the design of the study and by the covariate characteristics of the included patients in the analysis. The design refers to the number of subjects and their elementary designs, which means their dosage regimens and the allocation of their measurement times. Being able to predict the SE of the covariate effect estimates, in order to predict the ability to conclude on the significance and relevance of an effect is a key issue for the design of a clinical trial. For this reason, at design step, it is recommended to perform clinical trial simulation (CTS) to determine an adequate sample size or an adequate dispersion of the covariate to properly estimate covariate effects [6]. Nevertheless, this approach can be computationally expensive, and one aim of this work was to develop a method avoiding these computationally intense CTS for design evaluation through the use of the Fisher Information Matrix (FIM). This type of methodology is described in the general theory of optimal experimental design, which is applied to traditional non-linear models [7]. Indeed, according to the inequality of Rao-Cramer the variance of any unbiased estimator is bounded by the inverse of the FIM. Therefore, expected SE resulting from a design can be computed from it. In NLMEM framework there is no analytical solution for FIM computation, and the usual approach consist in a first order linearization of the structural model, around a mean of 0 for the random effects (FO method) or around individual realizations of the random effects (FOCE method) and then the FIM is computed as the FIM of the obtained Gaussian approximation [8]. This methodology has been shown to be appropriate for evaluating and optimizing design of experiments [9]. In this computation, the covariates are generally considered as being known for all the subjects, although accounting for their distributions, especially for continuous covariates is a priority for pharmaceutical companies in terms of clinical trial design and tools [10]. From the SE predicted by the FIM, the power of a parametric test to detect whether a covariate effect is statistically significant can be derived. Previous works have explored this computation for example including one discrete covariate and have shown that the trial design can be optimized to achieve a desired confidence level [11] [12]. These solutions have been implemented in *PFIM*, which was the first software tool proposed in 2001 for evaluating a design without using simulations. Especially, *PFIM* version 4.0 [13] includes evaluation methods for discrete covariates only while the latest version, the R package *PFIM 6*.*03* [14] (CRAN release March 2024) does not include covariates. Other software tools relying on this FIM based approach have been developed and handle both discrete and continuous covariates, such as PopED [15, 16] and NONMEM$DESIGN [17]. In PopED, covariates values are considered as known in FIM computation. Otherwise, to improve the computation, it is suggested in the user-guide to assume a distribution for the covariates and sample from it for each subject, then repeat the process and compute the average standard errors across the repetitions. NONMEM$DESIGN handles covariate similarly. The drawback of this assumption is the need to know the covariate values for each subject.

Furthermore, while the FIM gives asymptotic SE for covariate parameters, the quantity of interest is the ratio of change in PK parameters when covariate values change relative to a reference value. For classical covariate relationship models, an analytical solution allows to compute the CI on theses ratios from the value of the covariate parameters and its SE derived from the FIM, otherwise approximation such as the Delta method can be used. Thereafter, it is possible to predict the power of relevance test and to predict the CI on a forest plot, and thus evaluate whether the design leads to sufficient information to conclude to the relevance of covariates.

The goals of this work were thus to extend *PFIM6* to include both discrete and continuous covariates, to explore different approaches to handle continuous covariates and to predict SE of ratio, power of significance and relevance tests. To that purpose the computation of the linearized FIM was extended to the case with continuous covariates, comparing three methods to account for covariate distribution based on Monte-Carlo computation as already proposed [18]. The first one uses covariate vectors data, the second one independent distributions for the covariates, and the third one copulas, as the benefit of using copulas in pharmacometrics has already been demonstrated [**zwep2022virtual**]. The power of a clinical relevance test on covariate effect was computed analytically. These methods were implemented in a working version of *PFIM* 6 and are available in the Zenodo repository https://doi.org/10.5281/zenodo.13692989. Based on a simple PK model, simulations were performed to assess the accuracy of the proposed approaches. CTS and PFIM were compared in 4 scenarios, varying the sample size, the observation schedule and the IIV. Thereafter, this methodology was applied to a real example inspired from a population PK analysis of the drug cabozantinib [19] performed from 10 clinical trials, in healthy volunteers and patients with various cancer types The original Population PK model included 27 covariate relationships. The theoretical design was evaluated using the proposed methodology and the power of tests and the number of subjects that would have been required to achieve 80% power were calculated.

In Section 2, the notations and methods are detailed. In Section 3 we go through the simulation study, while section 4 describes the application to the real example. Finally global results are discussed in section 5

## 2 Methods

In this section we detail notations and methods on study design as well as NLMEM and on the FIM computation accounting for covariates. After that, derivation of SE and CI on ratios from the FIM are shown. Then computation for power of significance and relevance tests, and for number of subjects needed to achieve given levels of power are detailed.

### 2.1 Design and NLMEM

For one subject indexed by *i*, the elementary design *ξ*_*i*_ is defined by its number of observations *n*_*i*_, some designs variables 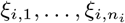 as observations time, doses, dosage regimens. The population design Ξ = {*N*, (*ξ*_1_, …, *ξ*_*N*_)} is composed of the number of subjects *N* and their respective elementary designs.

We denote *y*_*i*_ the *n*_*i*_-vector of observations for the *i*^*th*^ subject. Assuming Gaussian distribution for the observations, these can be modeled as given in equation (1), where *f* denotes the structural non-linear model. Thus the structural response depends on *ξ*_*i*_, the elementary design for subject *i* and on *θ*_*i*_ = *u* (*μ, β, z*_*i*_, *η*_*i*_), the *p*-vector of individual parameters. The latter is a function of *ν* = (*μ, β*) the vector of fixed effects, with *μ* the typical values and *β* the covariate effects, and of the vector of possibly transformed individual covariates *z*_*i*_ and individual random effects *η*_*i*_, where *η*_*i*_ ∼ 𝒩(0, Ω).

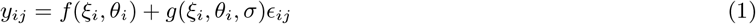

It is commonly assumed that the individual parameter can be transformed by a function *h* to a normal variable as given in equation (2).

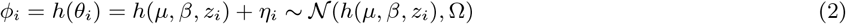

Moreover, it is usually assumed that each individual parameter *ϕ*_*il*_, *l* = 1, …, *p*, is a linear function of the possibly transformed covariates as given in equation (3), where *β*_*l*_ is a vector of size *C*, the number of covariates, and *z*_*pop*_ denotes the typical value of the (transformed) covariates in the population.

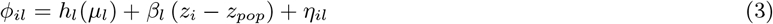

In PKPD, most of the parameters are modeled as log-normally distributed with additive covariate relationships on the log scale, i.e. 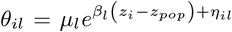 and *ϕ*_*il*_ = log *μ*_*l*_ + *β*_*l*_ (*z*_*i*_ − *z*_*pop*_) + *η*_*il*_, where *z*_*i*_ is the vector of (transformed) covariates.

The residual error *g*(*ξ*_*i*_, *θ*_*i*_, *σ*)*ϵ*_*i*_ is normally distributed with 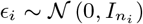. The function *g* models the residual error, and also depend on *ξ*_*i*_ and *θ*_*i*_, and on some parameters denoted *σ*.

The *P*-vector of population parameters is denoted Ψ ={*ν, λ*}, where *λ* contains the variance parameters (elements of Ω and of the residual error model *σ*).

### 2.2 Population Fisher Information Matrix computation with covariates

#### 2.2.1 Fisher Information Matrix

For a given elementary design *ξ*_*i*_ and a given vector of covariates *z*_*i*_, the expected elementary Fisher information is defined as the covariance matrix of the Fisher score (equation (4)) where *l*(*y*_*i*_; Ψ, *z*_*i*_, *ξ*_*i*_) is the likelihood of the vector of observations *y*_*i*_ for the population parameters Ψ, given the individual vector of covariates *z*_*i*_ and the elementary design *ξ*_*i*_ (equation (5)).

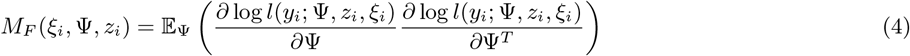

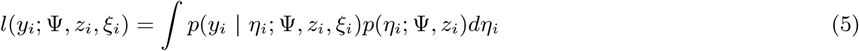

If the log-likelihood is twice differentiable, the FIM can be computed as minus the expectation of the Hessian of the log-likelihood (see equation (6)).

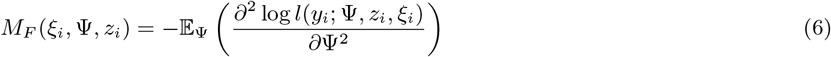

To calculate the standard error of a vector of population parameters, the standard method in NLMEM is to use the square root of the diagonal elements of the inverse of the FIM. According to the Rao-Cramer inequality, these values constitute the lower bound of the standard errors of any unbiased estimator of the parameters. We therefore assume in what follows that we are in asymptotic conditions and with an unbiased estimator such that: 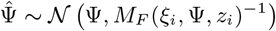.

#### 2.2.2 Calculation by linearization

Due to the non-linearity of the structural model *f* with respect to *θ*, there is no analytical expression for *l*(*y*_*i*_; Ψ, *z*_*i*_, *ξ*_*i*_). The expression for *M*_*F*_ (*ξ*_*i*_, Ψ, *z*_*i*_) are usually developed using a first-order Taylor expansion of the structural model around *ϕ*_*i*,0_ = *h*(*μ*_0_, *β*_0_, *z*_*i*_), a guess value of the fixed effects associated with covariates *z*_*i*_, and a zero-order expansion of *g* [8] as given in equation (7), where **J**_*θ*_*f* and **J**_*ϕ*_*h*^−1^ respectively denote the Jacobian matrix of *f* and *h*^−1^.

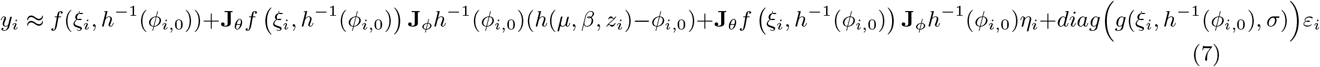

Observations are thus approximated to normal variables, with mean *E*_*i*_ (*ξ*_*i*_, *ϕ*_*i*,0_, *ν, z*_*i*_) and variance-covariance matrix *V*_*i*_ (*ξ*_*i*_, *ϕ*_*i*,0_, Ω, *σ*) given in equation (8).

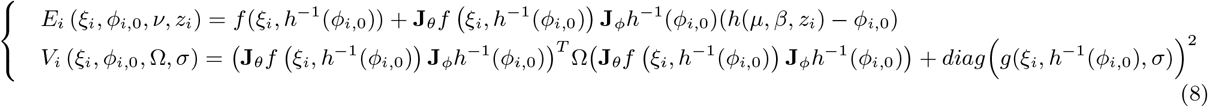

From this, the FIM is calculated as the FIM of the Gaussian vector obtained by approximation (see for example [18]).

#### 2.2.3 Calculation with covariates

In previous expressions, *M*_*F*_ (*ξ*_*i*_, Ψ, *z*_*i*_) is a function of *z*_*i*_. There are two main cases. First, if the covariate vector is known for each subject, *M*_*F*_ (*ξ*_*i*_, Ψ, *z*_*i*_) can be computed directly. Otherwise, *z*_*i*_ must be considered as a random variable and *M*_*F*_ (*ξ*_*i*_, Ψ) is compute as an integral with respect to *p*_*z*_, the distribution of the covariate vector as given in equation (9).

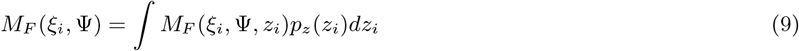

The population FIM of the design Ξ = {*ξ*_1_, …, *ξ*_*N*_}writes as the sum of the FIM of the elementary designs (see equation (10)).

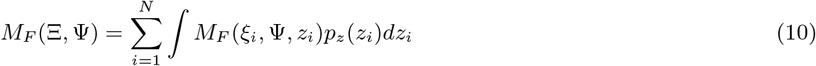

To compute *M*_*F*_ (Ξ, Ψ), a solution is to approximate it trough a Monte Carlo integration of the covariates as given in equation (11).

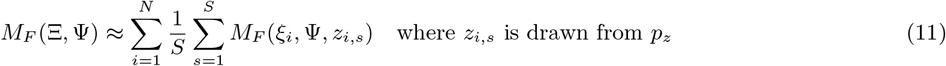

This method allows to account for the dependence structure between the covariates. However, it raises the question of how to draw the *z*_*i,s*_ used to calculate the integral. In this work, three methods were explored. The first one is non-parametric and consists in using an available dataset of covariate vectors, directly as a sample from the covariate distribution or as a discrete distribution and sample from it. This approach allows to account for the correlations between the covariates. Otherwise, vectors of covariates can be simulated using either a given probability distribution or a copula.

Copulas are cumulative distribution functions that capture the dependence structure between random variables and are therefore of interest to simulate correlated covariates. The benefits of using copulas for patients simulation in the field of pharmacometrics have already been demonstrated [20]. However, while the theory of copulas is well established for continuous variables, the same cannot be said for discrete variables. For a discrete vector, a copula cannot be uniquely identified and measures of dependence between variables become dependent on marginal laws. Methods for constructing copulas with discrete variables have been proposed (see for instance Geenens [21] for a review), but are beyond the scope of this work. In pharmacometrics, discrete covariates being in the vast majority of cases categorical covariates, we will be able to circumvent this problem by working with a copula by categories/combination of categories in the presence of several categorical covariates.

### 2.3 SE and ratio CI derivation from the FIM

Regarding covariate effects, the quantity of interest is often the ratio of change in primary parameters when covariate values change relative to a reference value *z*_*ref*_.

For categorical covariates, the expected ratio is calculated for the non-reference categories in relation to the reference category. For instance, for a binary covariate *z*_*ic*_ with reference *z*_*ic*_ = *z*_*pop,c*_ = 0, and non-reference *z*_*ic*_ = 1, the general formula for the ratio on the *l*^*th*^ parameter is given in equation (12).

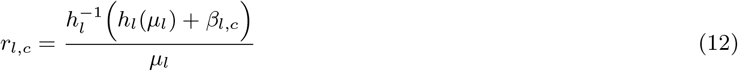

If *h* (*μ*) = *μ*, the ratio writes 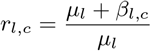 and if *h*(*μ*) = log *μ*_l_, it writes 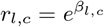.

For continuous covariates, the expected ratio is usually computed in the forest plot for the 10^*th*^ and 90^*th*^ percentiles of the covariate distribution, respectively denoted *P* 90 and *P* 10, in relation to its median denoted *P* 50, as given in equation (13);

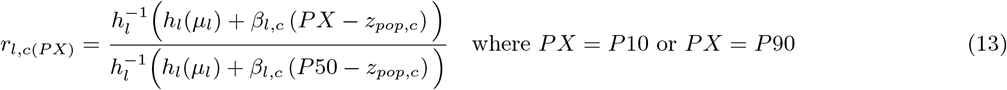

For instance, if *h* (*μ*) = *μ*, the ratio writes 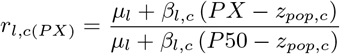 and if *h*(*μ*) = log *μ*_l_, it writes 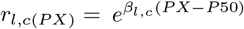.

The predicted CI at level 1 − *α* on a covariate parameter, assuming unbiased estimator, is *β ± q*_1 − *α/*2_*SE*_*β*_, where *q*_1− *α/*2_ denotes the 1 − *α/*2-quantile of the normal distribution. Therefore, when the ratio is a monotonic transformation of the covariate parameter only, the CI on the ratio can be easily derived. Otherwise, an approximation by the Delta method can be considered to also take into account the uncertainty on the typical value *μ*. For more complex cases, in particular to calculate the uncertainty of the ratio on secondary parameters such as AUC or Cmax, which depend on several model parameters, stochastic simulations can be carried out by sampling from a multivariate-normal centered on the parameter values and with variance-covariance matrix derived from the FIM. Details are for instance provided in Philipp et al. [22] and Guhl et al. [23].

In case of log-normally distributed parameter with additive covariate relationships on the log-scale, if *z*_*i,c*_ is binary: 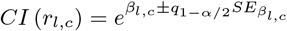, and if *z*_*i,c*_ is continuous: 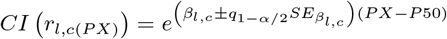.

### 2.4 Power of tests and Number of subjects needed

#### 2.4.1 Significance test

The statistical significance of a covariate parameter is assessed by a Wald test of comparison with null hypothesis *H*_0_ : *β*_*l,c*_ = 0 and alternative hypothesis *H*_1_ : *β*_*l,c*_ ≠ 0. The power of the significance test, *P*_*sign*_, at level 1 − *α* is given in equation (14). The detailed power calculation is presented in the Supplementary A.1.

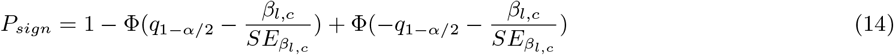

The number of subjects needed, *NSN*, to achieve a given power *P*_*sign*_ in significance test is given in equation (15).

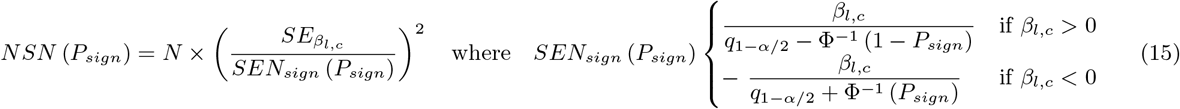

#### 2.4.2 Relevance test

The relevance of a covariate effect is assessed by a TOST procedure at level *α*, which is equivalent to the confidence interval approach with a 1 − 2*α* CI [5].

The null hypothesis is *H*_0_: “the covariate effect is not relevant”, i.e. *r*_*l,c*(*PX*)_ *∈* [*R*_*inf*_ ; *R*_*sup*_] while the alternative hypothesis is *H*_1_: “the covariate is relevant”, i.e. *r*_*l,c*(*PX*)_ *∉* [*R*_*inf*_ ; *R*_*sup*_]. This two sided null hypothesis can be split into two, respectively *H*_0,*inf*_ and *H*_0,*sup*_ :

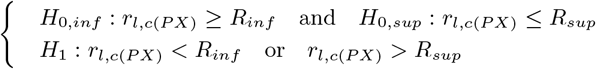

Thus *H*_0_ is not rejected unless neither *H*_0,*inf*_ nor *H*_0,*sup*_ is rejected.

For log-normally distributed parameter with additive covariate relationships on the log-scale, the ratio writes 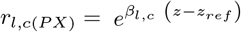, and at level 1 − *α*, the null hypothesis is rejected if 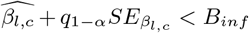 or if 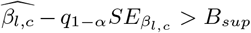 where depending on the sign of *z* − *z*_*ref*_, *B*_*sup*_ and *B*_*inf*_ equal 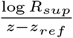 or 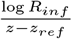.

The power is the probability under *H*_1_ to reject *H*_0_, it is given by the equation (16). The detailed calculations are presented in the Supplementary A.1.

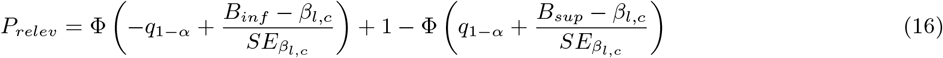

The number of subjects needed *NSN* to achieve a given power *P*_*relev*_ in relevance test is given in equation (17).

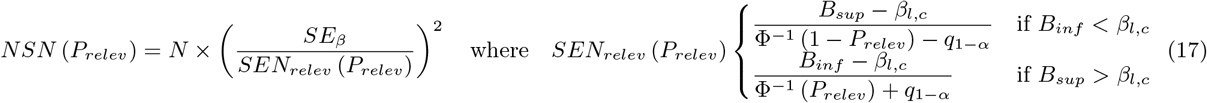

## 3 Evaluation by simulations

In this section, we implemented the methodology for computing the FIM using a simple PK model. Subsequently, we compared the predictions derived from this method with results from CTS. This comparison focuses on the uncertainty associated with covariate effects and their corresponding ratios, as well as the power of tests for both statistical significance and relevance.

### 3.1 Settings

#### 3.1.1 Data

The evaluation framework was inspired by a case study from the University of Maryland [24]. The data are from a double blind, parallel, single dose intravenous (IV) bolus trial, in which *N* = 100 patients were randomized equally in two arms receiving respectively 100 mg and 250 mg of drug. Concentration measurements from blood samples were collected at 16 times points (0.25, 0.5, 0.75, 1, 1.5, 2, 2.5, 3, 4, 6, 8, 10, 12, 16, 20 and 24 hours post-dose). This design is hereafter referred as Design 1. Covariates were Age, *Weight*, creatinine clearance (*CLCR*) and *Sex*, with a 0.82 correlation between *Weight* and *CLCR*. Only the covariate data were used in the following.

#### 3.1.2 Model

The PK model was a one compartment model with IV bolus and linear elimination, 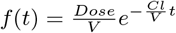, with two parameters: the clearance (*Cl*) and the volume of distribution (*V*). The residual error was modeled as a combined error model: *g* = *a* + *bf*. Continuous covariates *CLCR* and *Weight* were first log-transformed, and effects of log *CLCR* on *Cl* and log *Weight* and *Sex* on *V* were modeled as additive on the log-scale, as shown in equation (18) where 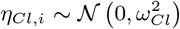 and 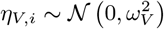 and with *Sex*_*i*_ = 0 if the *i*^*th*^ subject is a Male and 1 otherwise.

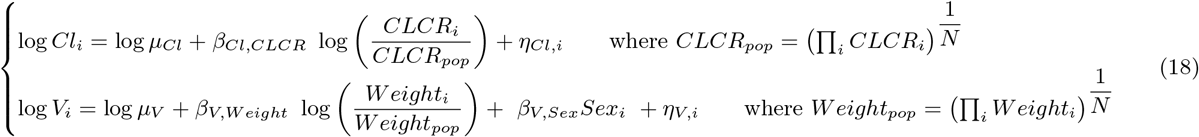

The PK parameter value used were the estimated values for a model without *Sex* effect on the initial dataset and are given in Table 1. *Sex* effect on *V* was arbitrarily set to *β*_*V,Sex*_ = − 0.35. The ratio values corresponding to the covariate parameters are also given in Table 1. It should be noted that all the covariate effects considered were statistically significant and clinically relevant.

**Table 1:** Evaluation - Population PK parameter values.

| Parameter | $\mu_{Cl}$ (L/h) | $\mu_V$ (L) | $\omega_{Cl}$ | $\omega_V$ | $a$ (mg/L) | $b$ | $\beta_{Cl,CLCR}$ | $\beta_{V,Weight}$ | $\beta_{V,Sex}$ |
| --- | --- | --- | --- | --- | --- | --- | --- | --- | --- |
| Value | 0.43 | 7.13 | 0.30 | 0.24 | 2.39 | 0.08 | 0.87 | 1.0 | -0.35 |
| Ratio | $r_{Cl,CLCR}$ (P10) | $r_{Cl,CLCR}$ (P90) | $r_{V,Weight}$ (P10) | $r_{V,Weight}$ (P90) | $r_{V,Sex}$ | | | | |
| Value | 0.62 | 1.66 | 0.65 | 1.44 | 0.70 |  |  |  |  |

$\mu$ .: refers to the typical value, $\beta$ .: covariate parameter, $\omega$ : standard deviation of the random effect, $r_{l,c}$ : ratio on the parameter $l$ for the covariate $c$

#### 3.1.3 Scenarios

Concentration evolution was studied in the reference scenario and in three other scenarios chosen to be more challenging. Those 4 different scenarios are obtained combining two design options and two variances options for random effects. Design 1 is the initial design of the study, and is composed of *N* = 100 subjects, divided equally between the 100 mg and 250 mg arms, with 16 sampling time points from 0.25 to 24h post dose. Design 2 is composed of *N* = 24 subjects only, also divided equally between the 100 mg and 250 mg arms, with 3 time points at 1, 4 and 12h post dose. For the variance, the first option, called True Omega keep the values *ω*_*Cl*_ and *ω*_*V*_ given in Table 1, while the second, called High Omega, doubled these standard deviations.

### 3.2 Prediction from FIM

The *PFIM* 6 source code has been extended in a working version to take account of both discrete and continuous covariates.

The FIM was computed by linearization of the structural model with first-order approximation and the three methods to handle covariates were considered. The first one used the 100 covariate vectors from the data: in the 4 scenarios, the FIM is computed with *S* = 100 by computing the elementary FIM for each of the vectors. The other two methods simulated *Z*_1_, …, *Z*_*S*_ with *S* = 1000 covariate vectors, using either independent distributions fitted on the data, namely independent Gaussian distributions for continuous covariates and Bernoulli distribution for discrete covariates, or two copulas fitted on the covariate table, one for each sex. Vine copula were fitted using the R package *rvinecopulib*0.6.3.1.1 [25] and the code made available by other researchers at https://github.com/vanhasseltlab/copula_vps [**zwep2022virtual**].

The Figure 4 given in appendix shows the fitted joint distribution of 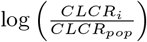 and 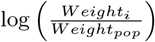 for Males and Females, with the Independent Gaussian approach and with Copulas. As expected, regarding marginal densities, the two methods give similar distributions while only Copulas can capture the correlation between the two covariates very precisely.

The predicted SE were computed as the square root of the diagonal elements of the FIM and used to computed the predicted NSE=RSE on the parameters: 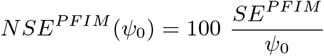. The predicted 90% CI on the ratios were also derived, as described in 2.3. The predicted power of significance test at level 95% and power of relevance test for the interval [0.80; 1.25] were calculated, as given in equations (14) and (16).

All other things being identical (i.e., same sampling scheme, same dosing and same covariate distributions), *PFIM* was also used to compute the power of comparison and relevance tests, for the simulation value *β*_*Cl,CLCR*_ = 0.87 (corresponding to the ratio *r*_*Cl,CLCR* (P10)_ = 0.62) as a function of *N*, for a smaller effect *β*_*Cl,CLCR*_ = 0.66 (*r*_*Cl,CLCR* (P10)_ = 0.70) and for a stronger effect *β*_*Cl,CLCR*_ = 1.3 (*r*_*Cl,CLCR* (P10)_ = 0.50).

### 3.3 Simulation

The same scenarios were explored by CTS in order to compare the estimates with the FIM predictions.

#### 3.3.1 Implementation

*S* = 200 datasets were simulated with R. For each subject, covariates were kept, and random effects on *Cl* and *V*, namely *η*_*Cl,i*_, *η*_*V,i*_, and the residual error *ϵ*_*i*_ were simulated.

#### 3.3.2 Estimations

Estimations were performed using SAEM algorithm implemented in Monolix 2023R1 [26], with default settings regarding auto-stop criteria, number of iterations, number of chains, and simulated annealing. Starting points were set to the simulation values for the base parameters and to 0 for the covariate effects. The expected FIM was computed within Monolix, by linearization around the Empirical Bayes Estimates (EBE) of the individual parameters.

The accuracy of parameter estimates was assessed by computing the relative estimation errors (*REE*) and by evaluating the Relative Bias (RB) and the Relative Root Mean Square Error (RRMSE) according to equation (19), where 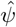 and *ψ*_0_ are the estimated and simulated parameters, respectively.

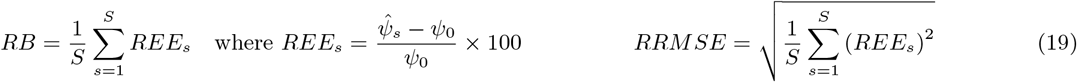

For each dataset, estimated SE, 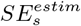, were calculated using the inverse of the expected FIM obtained by linearization.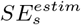 were normalized with respect to the true value *ψ*_0_ used for simulation: 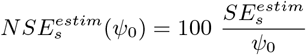.

The Empirical normalized SE across the datasets, *NSE*(*ψ*_0_)^*emp*^ were also computed as the mean of the normalized SE.

Estimated RSE, 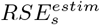, were computed: 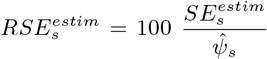 and the Empirical RSE 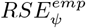 across the datasets were computed as the mean of the estimated RSE.

Ratio and their 90% CI were computed using the point estimates 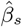 and the 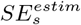.

Comparison test and relevance test were performed for each of the covariate effect. For the comparison test, the null hypothesis was rejected if 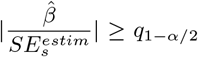 with *α* = 0.05. For the relevance test, the null hypothesis was rejected if the 90%CI on the ratio was outside [0.80; 1.25].

### 3.4 Evaluation methods

RSE predicted with PFIM were compared to median and quantiles of estimated NSE and RSE. Predicted CI on ratios were compared to median and quantiles of estimated bounds of CI on ratio. Predicted Powers were compared to mean and 95% CI on mean of estimated powers.

### 3.5 Results

#### 3.5.1 Estimation accuracy

In the 4 scenarios, all covariate parameter estimates were unbiased, see Figure 5 in the Supplementary, where *REE* boxplots were centered on 0. With Design 2 (i.e., fewer subjects and sparser observations), variance parameters had small negative bias (with True Omega, *RB*(*ω*_*Cl*_) = −20.1% and *RB*(*ω*_*V*_) = −9.99%; with High Omega, *RB*(*ω*_*Cl*_) = −11.5% and *RB*(*ω*_*V*_) = −9.31%).

#### 3.5.2 Uncertainty evaluation

Uncertainty was well predicted with PFIM, see Figure 6 in the Supplementary. Indeed, the three methods for handling covariates in the FIM gave similar results, as in all the scenarios and for almost all parameters, PFIM predictions were very close to empirical *NSE* and *RSE*.

With Design 1, for both True Omega and High Omega, SE were very well predicted by PFIM. Of note, with High Omega the empirical *RSE* for *β*_*V,SEX*_ was slightly larger that PFIM prediction and empirical *NSE*.

With Design 2, estimated *RSE* showed a large variability and because of few extreme values, the empirical *RSE* for some parameters were quite larger that the median *RSE* which remained close to the empirical *NSE*. With Design 2 and both True Omega and High Omega, SE were overpredicted for the residual error parameters compared to the empirical *NSE* but were smaller than the empirical *RSE*. For covariate parameters, PFIM predictions were in accordance with empirical *NSE* for the covariate effects. We can note that PFIM prediction for SE on *β*_*V,SEX*_ were slightly higher than the empirical *NSE* and the median *RSE*.

#### 3.5.3 Forest plots

Forest plots are shown on Figure 1, with PFIM predictions according to the three methods for handling covariates in green. The distributions of the estimated ratios are shown in red boxplots while the estimated bounds of the 90% CI on the ratios are shown in purple boxplots.

**Figure 1:**
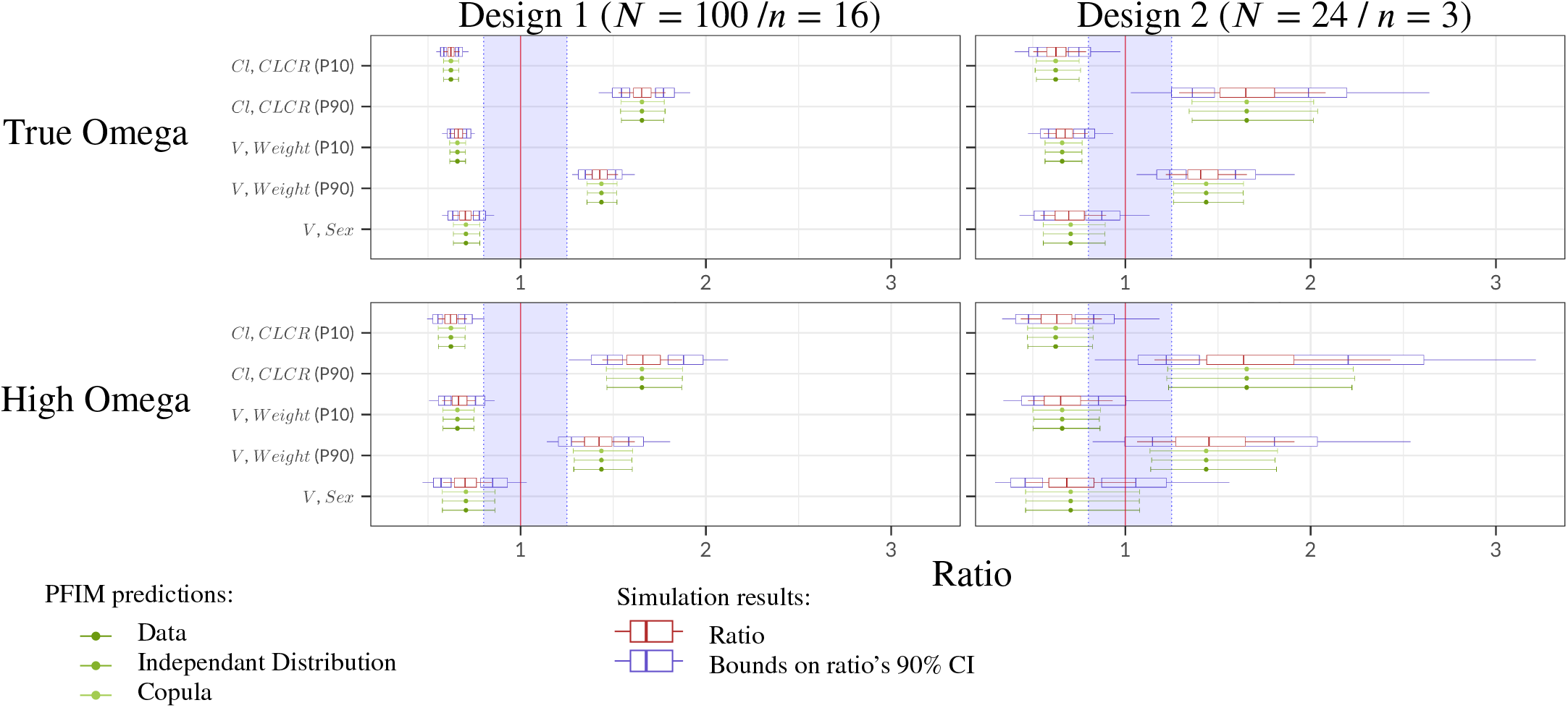
Evaluation - Forest Plots for the 4 scenarios: PFIM predictions using the three methods for handling covariates and simulation results across 200 datasets The red line at 1 corresponds to the reference line i.e., no change from the typical individual; the shaded area in blue represents the reference area of [0.80, 1.25]. The boxplot displays the median, the 25th and 75th percentiles, while the whiskers are 5th and 95th percentiles.

The confidence intervals for the ratios were overall well predicted for all the ratios, as the bounds of predicted CI were always very close to the median of the estimated bounds.

With Design 1 and True Omega, all the ratios were predicted relevant. When increasing IIV to High Omega, the CI of *Sex* effect on *V* crossed the [0.80; 1.25] area, so one cannot conclude to relevance.

In accordance with the slightly overprediction of SE on *β*_*V,Sex*_, it can be noted that with Design 2, the predicted CI on the ratio of *Sex* effect on *V* was larger than the CI formed by the respective median of the estimated lower and higher bounds of the 90%CI. In Design 2, the CI of the effect of the 90^*th*^ percentile of *CLCR* and *Weight*, respectively on *Cl* and *V* were slightly larger than the estimated CI.

With Design 2 and True Omega, only the CI of *Sex* effect on *V* crossed the [0.80; 1.25], therefore the effects of *CLCR* on *Cl* and *Weight* on *V* were predicted relevant for the two percentiles. When increasing IIV to High Omega, none of the parameter was predicted relevant.

#### 3.5.4 Power

##### Significance test

For the 4 scenarios, the predicted power for significance test derived from the SE computed from the FIM and the estimated power computed as the mean of the rejection of the null hypothesis across the datasets are shown in Table 2.

**Table 2:**
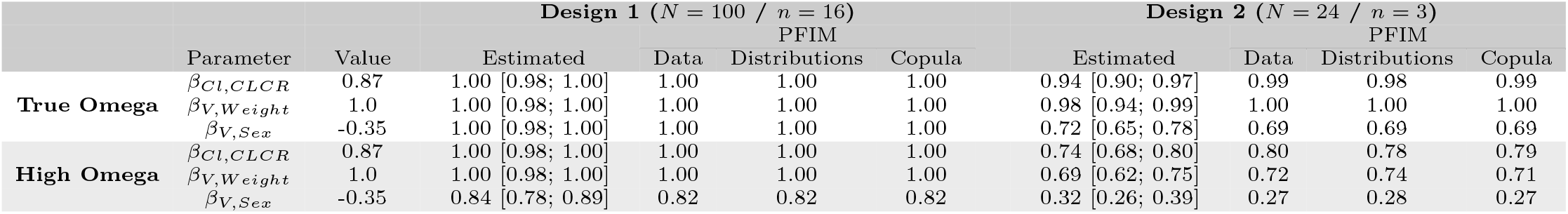
Evaluation - Power of significance test for the 4 scenarios: estimated power and its 95% CI across 200 datasets and PFIM prediction with the three methods for handling covariates.

First, as the SE were very close between the three methods for handling the covariates in the FIM computation, the powers were also the same for the three approaches.

With Design 1 and True Omega, comparison test on all the *β* parameters were predicted to 1 and estimated to 1 by CTS. Thus PFIM predictions were right.

With Design 1 and High Omega, power were very slightly overperdicted for *β*_*Cl,CLCR*_ and *β*_*V,Weight*_, with estimated 95% CI being respectively [0.90; 0.97] and [0.94; 0.99] and predictions being 0.99 and 1. For *β*_*V,Sex*_, the estimated power was 0.72 and FIM prediction was 0.69, thus lying in the estimated power’s 95% CI which was [0.65; 0.78].

With Design 2 (i.e., fewer subjects and sparser observations), PFIM prediction were very close to the estimated power and always within its 95% CI in case of both True Omega and High Omega.

##### Relevance test

For the 4 scenarios, the predicted power for relevance test derived from the SE computed from the FIM and the estimated power computed as the mean of the rejection of the null hypothesis across the datasets are shown in Table 3.

**Table 3:**
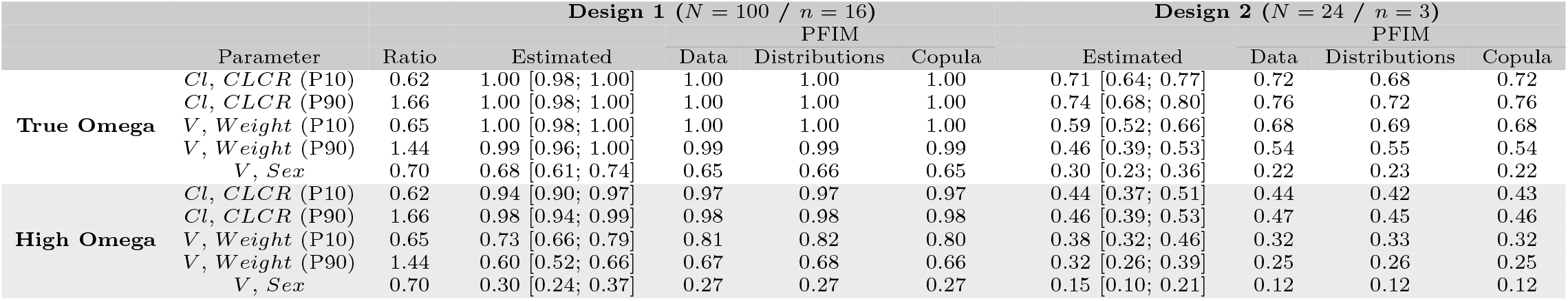
Evaluation - Power of relevance test for the interval [0.8, 1.25] for the 4 scenarios: estimated power and its 95% CI across 200 datasets and PFIM prediction with the three methods for handling covariates.

As for significance test, with Design 1 and True Omega, power of relevance tests were close to 1 and well predicted for effects of *CLCR* on *Cl* and *Weight* on *V*. For *Sex* effect on *V*, PFIM prediction 0.66 lied within the 95% CI of the estimated power [0.61; 0.74].

With Design 1 and High Omega, power of relevance tests were well predicted for effects of *CLCR* on *Cl* and slightly overpredicted for effect of *Weight* on *V* (respectively predicted at 0.68 with upper bound of the 95% CI at 0.66 for P10 and predicted at 0.54 with upper bound at 0.53 for P90). For *Sex* effect on *V*, PFIM prediction 0.22 was slightly below the 95% CI of the estimated power [0.23; 0.36].

Similarly, with Design 2 and True Omega, power of relevance tests were well predicted for effects of *CLCR* on *Cl* and *Sex* effect on *V* but slightly overpredicted for effect of *Weight* on *V* (respectively predicted at 0.81 with upper bound of the 95% CI at 0.79 for P10 and predicted at 0.67 with upper bound at 0.66 for P90).

With Design 2 and High Omega, power of relevance test were well predicted for all the parameters, just slightly underpredicted at 0.25 for *Weight* on *V* P90 while the lower bound of the observed CI was 0.26.

Overall, we can conclude that the FIM predictions were very satisfactory.

##### Evolution of power of test

Another important result from this methodology is being able to compute the power of tests depending on covariate effect magnitude, sample size, design or IIV. Thus is given on Figure 2 the power of significance test on *β*_*Cl,CLCR*_ and the power of relevance test for effect of *CLCR* (P10) on *Cl*, as function of *N* for different values of *β*_*Cl,CLCR*_, sampling schemes from Design 1 and 2 and IIV setting to True Omega or High Omega.

**Figure 2:**
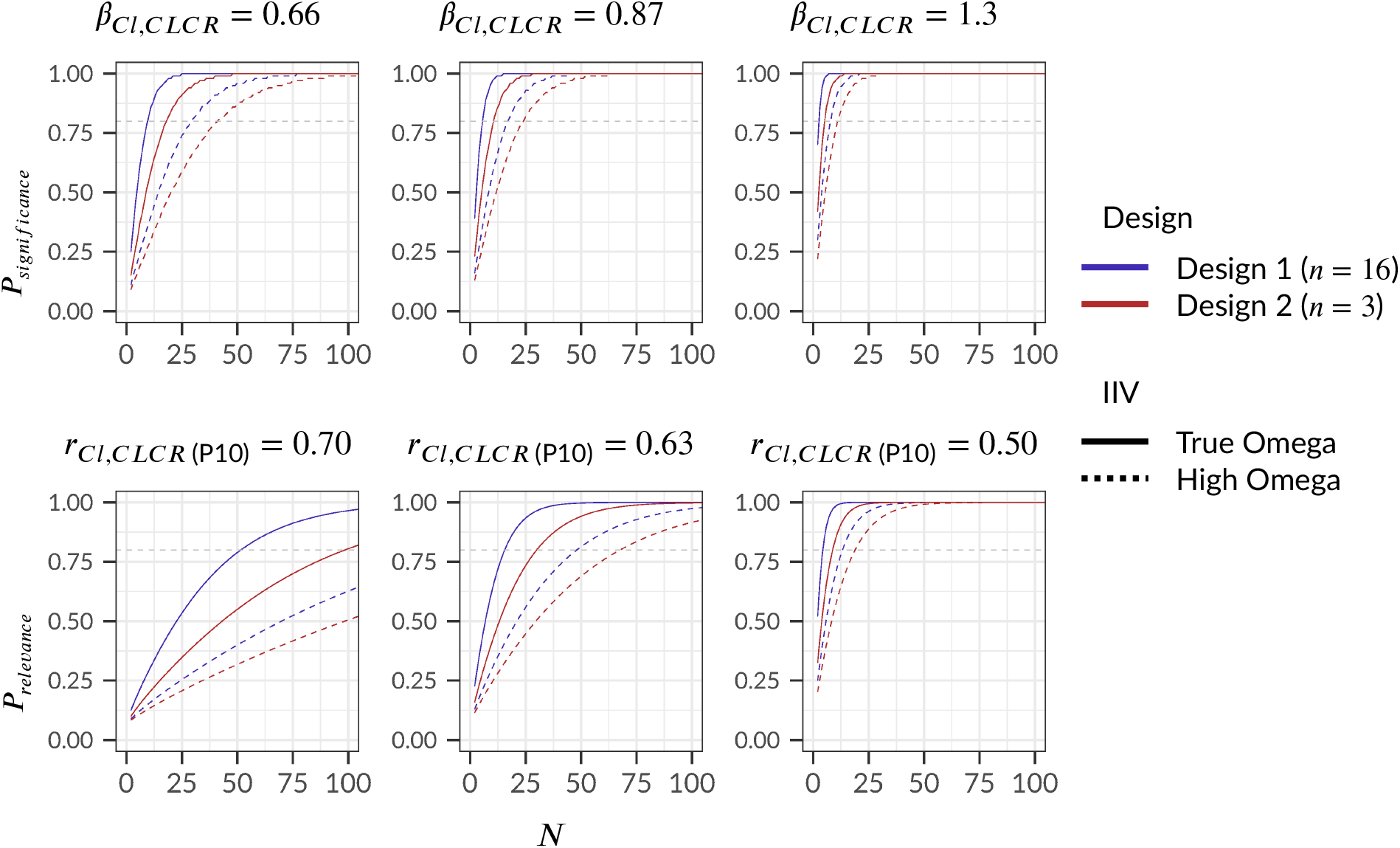
Evaluation - Predicted Power of significance test on *β*_*Cl,CLCR*_ and Power of relevance test for effect of *CLCR* (P10) on *Cl* for the interval [0.80; 1.25], as function of *N* and for different values of *β*_*Cl,CLCR*_: PFIM predictions using Data method.

The first foreseeable result shown by this plot is that the smaller the covariate effect, the smaller the powers of tests, which led to a higher number of subjects needed to achieve a given level of power. In addition, with sparser design (shown in red) the power of tests were always smaller than with rich design (shown in blue). The larger the IIV on parameters (dashed line), the lower the power of tests. We can also note that in this example, the variability had a greater impact than design, as switching from rich design with True Omega (blue plain line) to rich design with High Omega (blue dashed line) decreased power of both tests more than switching to sparse design with True Omega (red plain line).

## 4 Application to a population PK example of cabozantinib

In this section, the aim was to validate the method to compute the FIM for model with continuous and/or discrete covariates, in a more complex set up in terms of PK model and covariate structure.

This study was inspired from a real population PK analysis [19] performed on 10 clinical trials in healthy volunteers and patients with various cancer types receiving cabozantinib daily. The original data included 9510 quantifiable cabozantinib concentrations from 2023 subjects. The table of covariate vectors from this analysis were used in the present work.

### 4.1 Settings

#### 4.1.1 Covariate Data

The covariate data are pooled from the 10 clinical studies analysed in the original article. A summary is given in Table 4. Continuous covariates were Dose, Age, and Weight, while categorical covariates were Sex, Race, Population category and Formulation. It should be noted that certain categories are under-represented: for Race, only 10% of patients are Asian and only 3% Black and 2% from other race; and for pathology, only 10% of patients suffer from Metastatic medullary thyroid cancer (MTC) and 2% from Glioblastoma multiforme (GB). The baselines values from the 2023 subjects were used.

**Table 4:**
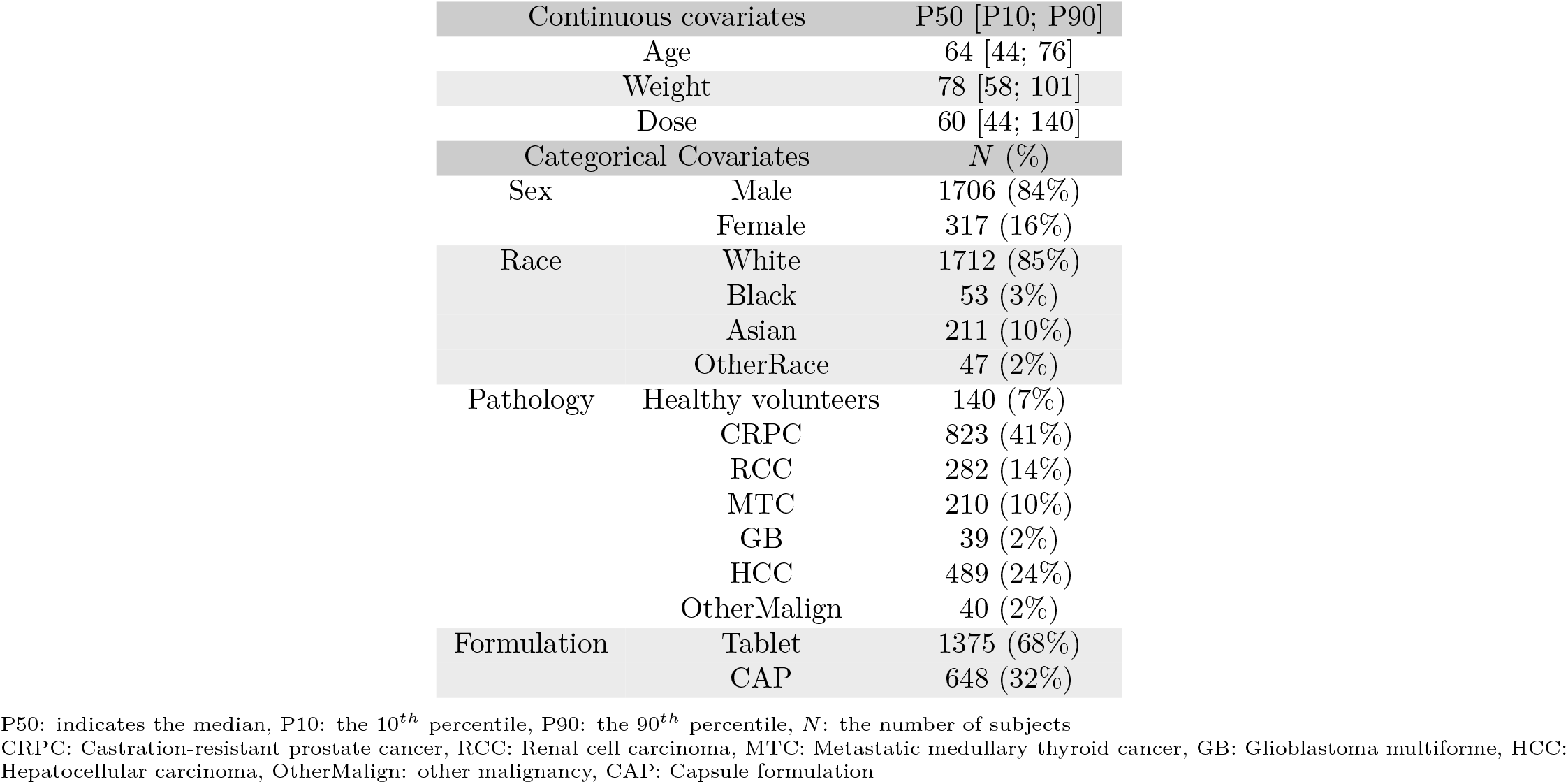
Application - Covariates summary, data pooled from the 10 clinical studies analysed in Nguyen et al.[19] combining 2023 subjects.

| Continuous covariates |  | P50 [P10; P90] |
| --- | --- | --- |
| Age |  | 64 [44; 76] |
| Weight |  | 78 [58; 101] |
| Dose |  | 60 [44; 140] |
| Categorical Covariates |  | N (%) |
| Sex | Male | 1706 (84%) |
|  | Female | 317 (16%) |
| Race | White | 1712 (85%) |
|  | Black | 53 (3%) |
|  | Asian | 211 (10%) |
|  | OtherRace | 47 (2%) |
| Pathology | Healthy volunteers | 140 (7%) |
|  | CRPC | 823 (41%) |
|  | RCC | 282 (14%) |
|  | MTC | 210 (10%) |
|  | GB | 39 (2%) |
|  | HCC | 489 (24%) |
|  | OtherMalign | 40 (2%) |
| Formulation | Tablet | 1375 (68%) |
|  | CAP | 648 (32%) |

P50: indicates the median, P10: the 10<sup>th</sup> percentile, P90: the 90<sup>th</sup> percentile, N: the number of subjects
CRPC: Castration-resistant prostate cancer, RCC: Renal cell carcinoma, MTC: Metastatic medullary thyroid cancer, GB: Glioblastoma multiforme, HCC: Hepatocellular carcinoma, OtherMalign: other malignancy, CAP: Capsule formulation

#### 4.1.2 Design

The studied design, given in Table 8 in the Supplementary, corresponded to the theoretical sampling designs of the 10 clinical trials analysed in the original article, associated with the number of included subjects and their doses. Three trials were phase I studies, with therefore few subjects (between 40 and 70) but with rich observations (up to 43 sampling points per subjects). Data also included two phase II studies with more balance between subjects and observations, and finally five phase III studies, with much more subjects (up to 498) but much sparser design with generally two to three samplings per subjects. In total, the studied design included 11621 observations from 2023 subjects.

#### 4.1.3 Model

For illustrative purposes and due to the current limitations of the *PFIM* package, the initial model [19] was simplified into a two-compartment model, with a first-order absorption discarding the dual absorption process, the presence of a lag time and a correlation between random effects. The analytical solution for the concentration in the central compartment is given in equation (20), where *Ka* is absorption rate constant, *Q/F* the apparent flow parameter between compartments, *V*_*c*_*/F* the apparent distribution volume of the central compartment and *V*_*p*_*/F* the apparent distribution volume of the peripheral compartment.

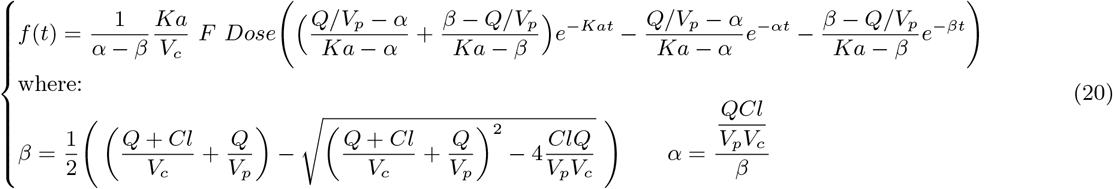

The model included log-normally distributed IIV on all the PK parameters; i.e., on *Ka, F, Cl, V*_*c*_, *Q* and *V*_*p*_. The residual variability is modeled using a proportional error model with variance *σ*^2^ *i*.*e*.*g* = *σf*.

All the covariate relationships of the initial model have been reproduced. It should be noted, however, that the initial covariate model was built without selection, notably with the same covariates affecting both *Cl* and *V*_*c*_, and that as a result, some covariate parameters values are very small and/or estimated with very wide confidence intervals leading to non statistically significant covariate effects. This is particularly true for the effects of categorical covariates for which the non-reference categories are very poorly represented (e.g. *MTC* patients represent 10% of the total population and *GB* only 2%).

Three continuous covariates were included in the model: *Dose, Age* and *Weight*. They were normalised by a reference value and log-transformed, as given in equation (21).

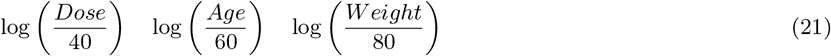

Tow binary covariates were included: *Sex* and Formulation, with the following definitions: *Sex* equals 0 if the subject is a Male (reference) and 1 otherwise and *CAP* equals 0 if the subject received a tablet formulation (reference), and 1 if the subject received a capsule formulation.

The categorical covariates included in the model were Race: White, Black, Asian or other race and Pathology: Healthy volunteers, Castration-resistant prostate cancer (*CRPC*), Renal cell carcinoma (*RCC*), Metastatic medullary thyroid cancer (*MTC*), Glioblastoma multiforme (*GB*), Hepatocellular carcinoma (*HCC*) and other malignancy (*OtherMalign*). These categorical covariates were converted into binary covariates by coding each category other than the reference as a separate binary variable, with 1 coding the corresponding category and 0 all others. Thus, for Race the reference was White and the categorical covariate was transformed to 3 binary covariates, respectively *Black, Asian* and *OtherRace*.

In the same way, for the Pathology, the reference was Healthy volunteers and 6 binary covariates were created: *CRPC, RCC, MTC, GB, HCC, OtherMalign*.

The reference was a healthy white Male subject with a body Weight of 80 kg, 60 years of Age, receiving a 40-mg tablet dose once daily.

The effects of the continuous covariates (*Dose, Age* and *Weight*) and of the binary covariates (*Sex, CAP, Black, Asian, OtherRace, CRPC, RCC, MTC, GB, HCC* and *OtherMalign*) were all additive on the logarithmic scale, as given by equation (22), where *β*_*θ,z*_ denotes the effect of covariate *z* on the parameter *θ*.

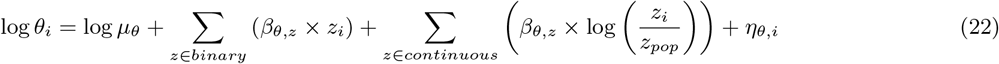

The model included effects of *Dose* and *CAP* on *Ka, CAP* on *F* and *Age, Weight, Sex, Black, Asian, OtherRace, CRPC, RCC, MTC, GB, HCC* and *OtherMalign* on both *Cl/F* and *V*_*c*_*/F*, which corresponded to 27 covariate effect parameters.

The values used were taken from [19] and are given in Table 5. We can already notice that some of the covariate effects were very small (e.g. *β*_*Cl/F,Asian*_ = −0.0668 corresponding to *r*_*Cl/F,Asian*_ = 0.94; *β*_*Cl/F,CRPC*_ = − 0.0115 corresponding to *r*_*Cl/F,Asian*_ = 0.99; *β*_*Cl/F,Age*_ = −0.157 corresponding to *r*_*Cl/F,Age* (P10)_ = 1.06 and *r*_*Cl/F,Age* (P90)_ = 0.97; *β*_*Cl/F,Weight*_ = 0.0393 corresponding to *r*_*Cl/F,Weight* (P10)_ = 1.01 and *r*_*Cl/F,Weight* (P90)_ = 0.99; 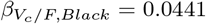 corresponding to 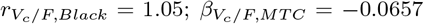 corresponding to 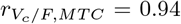 and 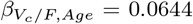 corresponding to 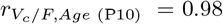 and 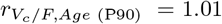. In addition, only a few relationships were relevant, in the sense that their ratio was outside [0.80; 1.25], those relationships were the effects of *CAP* on *Ka*, both *Dose* (P10) and (P90) on *Ka, Sex* on *Cl/F, MTC* on *Cl/F, Asian* on *V*_*c*_*/F, CRPC* on *V*_*c*_*/F, GB* on *V*_*c*_*/F, OtherMalign* on *V*_*c*_*/F* and both *Weight* (P10) and (P90) on *V*_*c*_*/F*, i.e. 11 ratios out of 32.

**Table 5:**
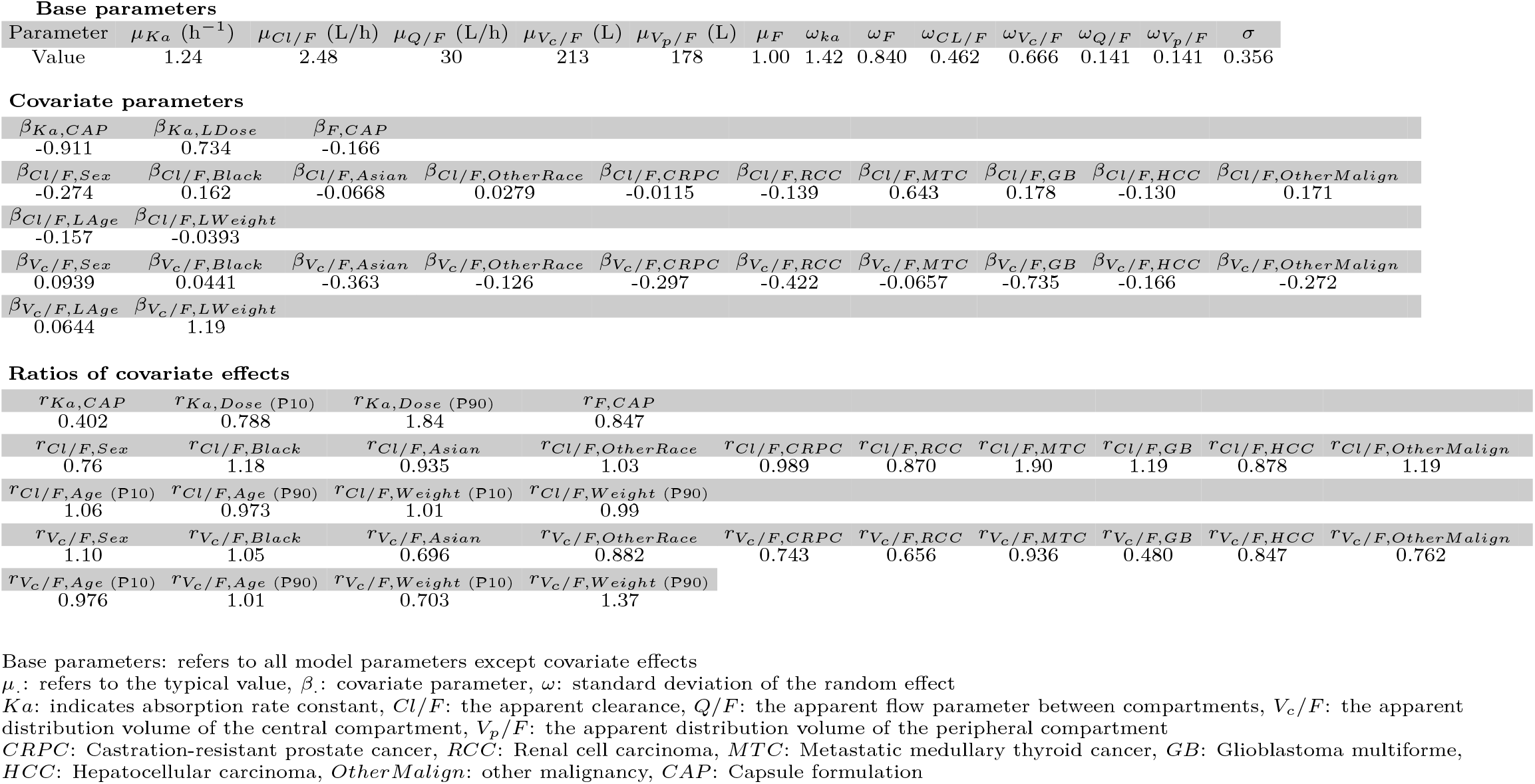
Application - Parameter Values taken from Nguyen et al.[19] and used in the present work.

### 4.2 Prediction from FIM

Because the three methods for handling covariates have shown similar performances in the first simulation study (see Section 3), only the Data method was kept. Thus, covariate were taken into account in the FIM computation through the covariate dataset: for each study and each dose (i.e., for each elementary design), the covariate vectors of the subjects corresponding to this elementary design were used.

As previously, from the predicted SE returned by *PFIM*, predicted NSE=RSE on the parameters were computed. Afterwards, predicted 90% CI on the ratios were derived, such as predicted powers of significance and relevance tests for the range [0.80; 1.25]. The number of subjects required to achieve 80% power, in the comparison test and the relevance test respectively, was calculated when the current predicted power was greater than 50%. These *NSN* hold as long as all other things remains identical, i.e., with the same observation points, the same dosing schemes and the same covariate distributions within the trial population.

### 4.3 Simulation

The same scenarios was explored by CTS in order to compare the estimates with the FIM predictions. *S* = 200 datasets were simulated with the R API of Simulix [27] and using the initial covariate dataset.

#### 4.3.1 Estimations

Estimations were performed using SAEM algorithm implemented in Monolix 2023R1 [26], with default settings regarding auto-stop criteria, number of iterations, and Simulated annealing; and 5 Markov chains. The starting point were set to the parameter values used for the simulation for the *μ, ω* and *σ*, and to 0 for the covariate effects *β. μ*_*F*_ was fixed to 1, and IIV on *Q* and *V*_*p*_ were fixed to *ω* = 0.141.

The accuracy of parameter estimates was assessed by computing the REE, the RB and the RRMSE.

As detailed in the section 4, for each dataset, estimated *NSE* and *RSE* were computed for all the parameters and the empirical *NSE* and *RSE* were secondly derived.

For each covariate parameter, significance test was performed through a Wald test of comparison with *α* = 0.05. Ratio and their 90% CI were computed and relevance tests are performed for the interval [0.80; 1.25].

#### 4.3.2 Evaluation methods

RSE predicted with PFIM were compared to median and quantiles of estimated NSE and RSE. Predicted CI on ratios were compared to median and quantiles of estimated bounds of CI on ratio. Predicted Powers were compared to mean and 95% CI on mean of estimated powers.

### 4.4 Results

#### 4.4.1 Estimation accuracy

The distribution of REE for base parameters and ratios on covariate effects are given in the Supplementary, respectively in Figures 7 and 8 and in Tables 9 and 10 assessing that parameters were overall accurately and precisely estimated.

Of note, some *RRMSE* were very large for covariate parameters: over 20% and up to more than 800% for very small *β* (e.g. *β*_*CL,CRPC*_ = −0.0115, *RRMSE* = 808%) and some biases over 10%, but one again for very small *β* (e.g. *β*_*CL,CRPC*_ = −0.0115, *RB* = −54.6%).

#### 4.4.2 Uncertainty evaluation

Uncertainty was well predicted with PFIM, see Figure 9 in the Supplementary for base parameters and 10 for covariate parameters. For base parameters, once again the Data method for handling covariates in the FIM gave SE predictions close to empirical *NSE* and *RSE*, except for *μ*_*Q*_, for which the predicted SE was 13.4% while the empirical *NSE* and *RSE* were 10.3%.

For covariate parameters, we can first notice at first glance that the SE, both predicted and estimated, were on the whole much larger than in the previous example, particularly for the effects of discrete covariates, some categories of which were poorly represented in the dataset (e.g. Asian, OtherRace). Indeed, for 12 covariate parameters out of 27 SE were above 100 %, even reaching more than 800% for 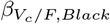 and *β*_*Cl/F,CRCP*_, recalling that those effects were very small, 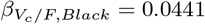 and *β*_*Cl/F,CRCP*_ = −0.0115 and that they were only 53 Black subjects (3%). In addition, there was a large uncertainty in empirical *RSE* and for 21 parameters out of 27, the empirical *RSE* was much larger than the median of estimated *RSE*. This is because the covariate parameters can be estimated very close to zero, which leads to a very high quantity when they are used as the denominator in a calculation. Nonetheless, for all the covariate parameters, PFIM prediction remained very close to the empirical *NSE*, and if there were differences, the orders of magnitude were always the same.

#### 4.4.3 Forest plots

Forest plots are shown on Figure 3, with PFIM predictions according to the Data method for handling covariates in green. The distributions of the estimated ratios are shown in red boxplots while the estimated bounds of the 90% CI on the ratios are shown in purple boxplots.

**Figure 3:**
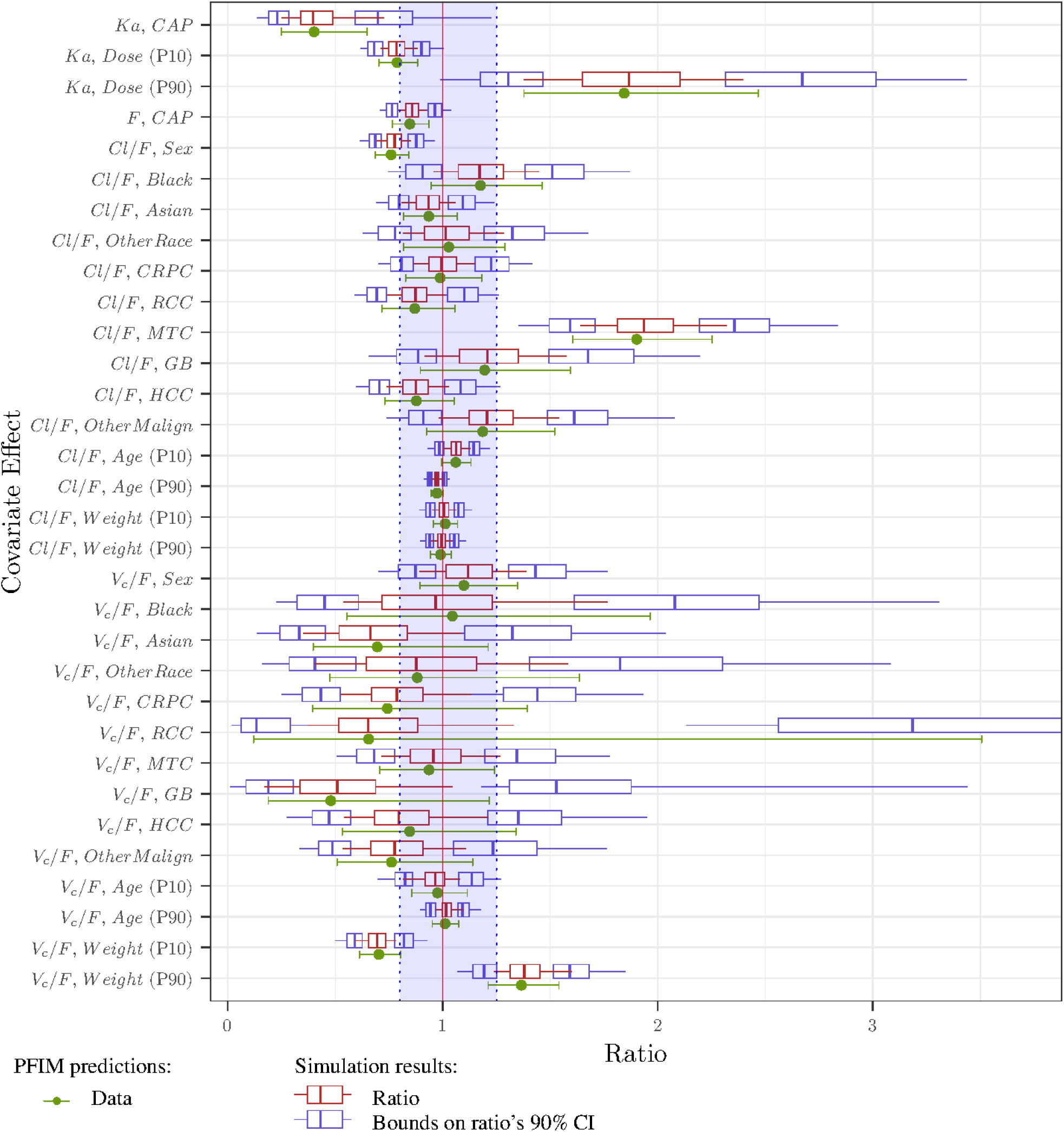
Application - Forest Plot: Ratios and their Confidence intervals: PFIM predictions using Data method for handling covariate and simulation results across 200 datasets The red line at 1 corresponds to the reference line i.e., no change from the typical individual; the shaded area in blue represents the reference area of [0.80, 1.25]. The boxplot displays the median, the 25th and 75th percentiles, while the whiskers are 5th and 95th percentiles.

As in the previous example, confidence intervals for the ratios were overall well predicted, even though there was an overall tendency for the upper bound of the interval to be slightly underpredicted. This can be explained if the covariate parameter was estimated with a slight bias leading to a slight bias on the ratio (which can be seen on the forest plot) and if 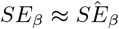. Then multiplying the ratio with 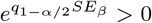 for computing the upper bound of the confidence interval inflated the bias between the predicted bound and the estimated bound. For instance, for effect of *GB* on *Cl/F*, bias on *β*_*Cl/F,GB*_ was 7.73% and FIM prediction for the SE was very close to the median RSE which led to a higher estimated bound for the CI on the ratio.

On the contrary, for *V*_*c*_*/F, RCC*, the predicted upper bound on the CI was higher than the estimated median because of the difference between the predicted SE 242% higher than the median *RSE*: 192%. However, for such large uncertainties, this was still equivalent.

In this example, with the given design, very few covariate effects were relevant for the interval [0.80, 1.25]. According to PFIM prediction, the effects of *CAP* on *Ka*; *Dose* (P90) on *Ka*, and *MTC* on *Cl/F* were relevant. This also shows that the covariate may be relevant for certain percentiles only.

#### 4.4.4 Power

##### Significance test

The predicted power for significance test derived from the SE computed from the FIM and the estimated power are shown in Table 6. PFIM predictions were very good, as lying in the 95% CI of the estimated power for 22 parameters out of 27. For *β*_*Cl/F,OtherRace*_ and 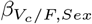, the prediction was slightly below the CI; for 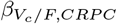 and 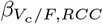 the prediction was slightly above while for 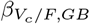 it was above: 0.25 against [0; 0.03]. This results comes from the bias in the ratio visible on the forest plot were the median of the estimated ratio is higher than the true value, and from the SE that was slightly under predicted: 77.0%, compared to empirical *NSE* 89.2% and median *RSE* 82.7%. However, it should be remembered that only 39 subjects (2%) were GB, which explains the difficulties in estimating this parameter.

**Table 6:** Application - Significance test: estimated power (with 95% CI) across 200 datasets, each including 2023 subjects, PFIM predicted power and number of subjects needed *NSN* to achieve a 80% power using the Data method.

| Parameter | Value | Estimated | PFIM | $NSN$ |
| --- | --- | --- | --- | --- |
| $\beta_{Ka,CAP}$ | -0.911 | 0.88 [0.83; 0.92] | 0.88 | 1 612 |
| $\beta_{Ka,Dose}$ | 0.734 | 0.94 [0.90; 0.97] | 0.93 | 1 331 |
| $\beta_{F,CAP}$ | -0.166 | 0.78 [0.72; 0.84] | 0.79 | 2 090 |
| $\beta_{Cl/F,Sex}$ | -0.274 | 0.99 [0.96; 1.00] | 0.99 | 845 |
| $\beta_{Cl/F,Black}$ | 0.162 | 0.23 [0.18; 0.30] | 0.23 | |
| $\beta_{Cl/F,Asian}$ | -0.0668 | 0.18 [0.13; 0.24] | 0.13 | |
| $\beta_{Cl/F,OtherRace}$ | 0.0279 | 0.07 [0.04; 0.12] | 0.05 | |
| $\beta_{Cl/F,CRPC}$ | -0.0115 | 0.03 [0.01; 0.06] | 0.05 | |
| $\beta_{Cl/F,RCC}$ | -0.139 | 0.20 [0.14; 0.26] | 0.22 | |
| $\beta_{Cl/F,MTC}$ | 0.643 | 1.00 [0.98; 1.00] | 1.00 | 409 |
| $\beta_{Cl/F,GB}$ | 0.178 | 0.22 [0.17; 0.29] | 0.17 | |
| $\beta_{Cl/F,HCC}$ | -0.13 | 0.22 [0.16; 0.28] | 0.22 | |
| $\beta_{Cl/F,OtherMalign}$ | 0.171 | 0.23 [0.18; 0.30] | 0.20 | |
| $\beta_{Cl/F,Age}$ | -0.157 | 0.30 [0.24; 0.37] | 0.32 | |
| $\beta_{Cl/F,Weight}$ | -0.0393 | 0.05 [0.02; 0.09] | 0.06 | |
| $\beta_{Vc/F,Sex}$ | 0.0939 | 0.18 [0.13; 0.25] | 0.12 | |
| $\beta_{Vc/F,Black}$ | 0.0441 | 0.06 [0.03; 0.10] | 0.05 | |
| $\beta_{Vc/F,Asian}$ | -0.363 | 0.14 [0.09; 0.19] | 0.19 | |
| $\beta_{Vc/F,OtherRace}$ | -0.126 | 0.09 [0.05; 0.14] | 0.06 | |
| $\beta_{Vc/F,CRPC}$ | -0.297 | 0.05 [0.02; 0.09] | 0.12 | |
| $\beta_{Vc/F,RCC}$ | -0.422 | 0.01 [0.00; 0.04] | 0.07 | |
| $\beta_{Vc/F,MTC}$ | -0.0657 | 0.06 [0.03; 0.10] | 0.07 | |
| $\beta_{Vc/F,GB}$ | -0.735 | 0.00 [0.00; 0.03] | 0.25 | |
| $\beta_{Vc/F,HCC}$ | -0.166 | 0.06 [0.03; 0.10] | 0.09 | |
| $\beta_{Vc/F,OtherMalign}$ | -0.272 | 0.18 [0.13; 0.24] | 0.20 | |
| $\beta_{Vc/F,Age}$ | 0.0644 | 0.08 [0.05; 0.13] | 0.06 | |
| $\beta_{Vc/F,Weight}$ | 1.19 | 1.00 [0.98; 1.00] | 0.99 | 870 |

CRPC: indicates Castration-resistant prostate cancer, RCC: Renal cell carcinoma, MTC: Metastatic medullary thyroid cancer, GB: Glioblastoma multiforme, HCC: Hepatocellular carcinoma, OtherMalign: other malignancy, CAP: Capsule formulation

The *NSN* to achieve a power of 80% was computed for covariate parameters for which the power of significance was already above 50% with the 2023 subjects. Only 6 covariate parameters matched this criteria: *β*_*Ka,CAP*_, *β*_*Ka,Dose*_, *β*_*F,CAP*_, *β*_*Cl/F,Sex*_, *β*_*Cl,MTC*_ and 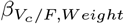, with respective powers even above 80%, therefore the *NSN* to reach a power of 80% were smaller than 2023. All other things being identical (i.e., same sampling scheme, same dosing and same covariate distributions), for instance only 409 subjects would have been enough to conclude statistical significance for *β*_*Cl/F,MTC*_. As 10% of the initial population were MTC patients, this would roughly correspond to 41 MTC patients and 369 patients from other categories, respecting their initial proportions.

##### Relevance test

The predicted power for relevance test derived from the SE computed from the FIM and the estimated power are shown in Table 7. PFIM predictions were overall slightly higher than what was really observed.

**Table 7:** Application - Relevance test: estimated power (with 95% CI) across 200 datasets, each including 2023 subjects, PFIM predicted power and number of subjects needed *NSN* to achieve a 80% power using the Data method.

| Parameter | Ratio | Estimated | PFIM | $NSN$ |
| --- | --- | --- | --- | --- |
| $Ka, CAP$ | 0.40 | 0.66 [0.58; 0.72] | 0.77 | 2 228 |
| $Ka, Dose$ (P10) | 0.79 | 0.02 [0.01; 0.05] | 0.08 | |
| $Ka, Dose$ (P90) | 1.84 | 0.62 [0.55; 0.69] | 0.71 | 2 598 |
| $F, CAP$ | 0.85 | 0.00 [0.00; 0.02] | 0.00 | |
| $Cl/F, Sex$ | 0.76 | 0.09 [0.05; 0.14] | 0.20 | |
| $Cl/F, Black$ | 1.18 | 0.01 [0.00; 0.04] | 0.02 | |
| $Cl/F, Asian$ | 0.94 | 0.00 [0.00; 0.02] | 0.00 | |
| $Cl/F, OtherRace$ | 1.03 | 0.01 [0.00; 0.04] | 0.00 | |
| $Cl/F, CRPC$ | 0.99 | 0.00 [0.00; 0.02] | 0.00 | |
| $Cl/F, RCC$ | 0.87 | 0.00 [0.00; 0.02] | 0.01 | |
| $Cl/F, MTC$ | 1.90 | 1.00 [0.97; 1.00] | 0.99 | 756 |
| $Cl/F, GB$ | 1.19 | 0.01 [0.00; 0.04] | 0.03 | |
| $Cl/F, HCC$ | 0.88 | 0.00 [0.00; 0.02] | 0.01 | |
| $Cl/F, OtherMalign$ | 1.19 | 0.01 [0.00; 0.04] | 0.02 | |
| $Cl/F, Age$ (P10) | 1.06 | 0.00 [0.00; 0.02] | 0.00 | |
| $Cl/F, Age$ (P90) | 0.97 | 0.00 [0.00; 0.02] | 0.00 | |
| $Cl/F, Weight$ (P10) | 1.01 | 0.00 [0.00; 0.02] | 0.00 | |
| $Cl/F, Weight$ (P90) | 0.99 | 0.00 [0.00; 0.02] | 0.00 | |
| $V_c/F, Sex$ | 1.10 | 0.00 [0.00; 0.03] | 0.00 | |
| $V_c/F, Black$ | 1.05 | 0.01 [0.00; 0.04] | 0.03 | |
| $V_c/F, Asian$ | 0.70 | 0.00 [0.00; 0.03] | 0.11 | |
| $V_c/F, OtherRace$ | 0.88 | 0.04 [0.02; 0.08] | 0.03 | |
| $V_c/F, CRPC$ | 0.74 | 0.00 [0.00; 0.03] | 0.07 | |
| $V_c/F, RCC$ | 0.66 | 0.00 [0.00; 0.02] | 0.08 | |
| $V_c/F, MTC$ | 0.94 | 0.00 [0.00; 0.03] | 0.01 | |
| $V_c/F, GB$ | 0.48 | 0.00 [0.00; 0.02] | 0.23 | |
| $V_c/F, HCC$ | 0.85 | 0.01 [0.00; 0.04] | 0.03 | |
| $V_c/F, OtherMalign$ | 0.76 | 0.03 [0.01; 0.06] | 0.07 | |
| $V_c/F, Age$ (P10) | 0.98 | 0.00 [0.00; 0.02] | 0.00 | |
| $V_c/F, Age$ (P90) | 1.01 | 0.00 [0.00; 0.02] | 0.00 | |
| $V_c/F, Weight$ (P10) | 0.70 | 0.38 [0.31; 0.45] | 0.47 | |
| $V_c/F, Weight$ (P90) | 1.37 | 0.26 [0.20; 0.32] | 0.33 | |

P10: indicates the 10<sup>th</sup> percentile, P90: the 90<sup>th</sup> percentile
CRPC: Castration-resistant prostate cancer, RCC: Renal cell carcinoma, MTC: Metastatic medullary thyroid cancer, GB: Glioblastoma multiforme, HCC: Hepatocellular carcinoma, OtherMalign: other malignancy, CAP: Capsule formulation

The *NSN* to achieve a power of 80% was computed for the ratios for which the power of relevance was already above 50% with the 2023 subjects, i.e., the effects of *CAP* on *Ka*; *Dose* (P90) on *Ka*, and *MTC* on *Cl/F*.

For the effects of *CAP* and *Dose* (P90) on *Ka* relevance power were respectively 77% and 78% and the *NSN* to reach 80% were respectively 2 228 and 2 598. Thus, all other things being identical, just over 600 additional subjects would have been needed to conclude that *Ka, CAP* and *Ka* and *Dose* (P90) were relevant, with a power of 80%. *MTC* having a strong effect on *Cl/F* (*r*_*Cl/F,MTC*_ = 1.90), the power of relevance was 99% and all other things being identical the level 80% would have been reached for only 756 subjects (including 10% of *MTC*).

## 5 Discussion

In this paper, we extended FIM computation accounting for continuous covariates. in the context of NLMEM. To that purpose, we introduced three methods for taking covariates into account when computing the FIM, and implemented them in a working version of the R package *PFIM*. This implementation is available (Zenodo repository https://doi.org/10.5281/zenodo.13692989) and could be used by others for further exploration. These methods are all based on Monte Carlo integration, but the first one relies on leveraging a covariate dataset directly, while the second one simulates from independent distribution and the last one simulates from copulas. For the latter, we used the code developed by other authors [**zwep2022virtual**] and made available online: https://github.com/vanhasseltlab/copula_vps. We showed that overall *PFIM* accurately predicts uncertainty on covariate effects and consequently the power of both significance and relevance tests. This method can then be used to display these predictions in the form of a forest plot for visual communication. We also used this approach to quantitatively describe that the power of both significance and relevance tests decrease when reducing the number of subjects or reducing the number of observations or increasing the IIV. The results of the three methods for handling covariates in FIM computation showed very similar performances in our simulation study, but their respective impact should be further explored in complex cases. Especially, if the covariates are highly correlated, we can expect the calculation using independent distributions to perform less well. In addition, we have chosen to propose an approach that fits independent covariates with Gaussian distributions rather than using a multivariate normal distribution to explore, using copulas, whether correlations play a role in this case. Nevertheless, we can imagine an intermediate method using a multivariate Gaussian, which are easier to interpret than Copulas.

The method was utilized in a practical example based on a population PK analysis of the drug cabozantinib, conducted across 10 clinical trials involving healthy volunteers and patients with various types of cancer. With its 27 relationships and many very low-value covariate parameters, this application represents, unlike the evaluation example, a case where there is a real challenge in determining the relevant effects. The proposed methodology was used to evaluate the theoretical design, computing the power of tests and the number of subjects needed to achieve 80% power. We found that despite the large number of relationships between covariates, some of which were not significant, and with certain covariates poorly represented in the data, PFIM was able to predict uncertainty very well and was very close to the CTS results. It is therefore an appropriate approach for rapidly computing the powers and the number of subjects required to achieve given powers, even in more complex contexts.

A limitation of the proposed methodology is that there is no consensus on how to take uncertainty into account when building forest plots, and simulations-based procedures often account for uncertainty in based parameters which is not the case here. Sometimes, the IIV is also considered in the simulations [2], even if some authors suggest not doing so in order to avoid misunderstanding [28]. Therefore, before using this method to predict forest plot, or comparing it with simulations, the randomness to be accounted for needs to be clearly defined.

In addition, the calculation of the confidence interval on the ratio is exact only in certain particular cases, otherwise an approximation, for example with the delta method, would be necessary. Moreover, our method is also less straightforward for deriving ratios and CI for secondary parameters such as Cmax or AUC and would require stochastic computations.

The main limitation of our methodology is the a priori knowledge of covariable distributions to compute the FIM. Indeed, without their distribution in the target population, the design evaluation cannot be performed. Nevertheless, the main use of this approach may be to design a phase III study based on phase II results, and at this stage of pharmaceutical development, expert usually have clues about the covariate distribution in the targeted population. In addition, if methods based on provided distributions is chosen, data are not mandatory as distributions can for instance be chosen according to expert opinion. In the same way, as copula do not require data, we can imagine using copulas from other projects or from other sources but corresponding to the target population of the trial. However, problems arise in the presence of discrete covariates, since the theory of copulas has not yet been fully extended to them, especially for non-ordered categorical data [29]. However, this is an active research area in pharmacometrics, and dedicated research and R package were recently published [29], it is therefore likely that solutions will be proposed in the near future.

Finally, these methods save a considerable amount of time compared with simulations, enabling a wider range of designs to be evaluated. Therefore, this tool can be used to study changes in the power of tests as a function of sample size, IIV, effect size or sampling design before conducting a trial to help in decision making. The speed of calculation could be further increased, for example by using efficient integration schemes (e.g. Quasi-Monte Carlo methods) or by deriving a analytical formula of the linearized FIM in the case where the continuous covariates follow a multivariate Gaussian distribution. These promising results suggest that the method should be more widely implemented in the *PFIM* package. In addition, this work opens the prospect of optimizing the distribution of covariates in clinical trials with the aim of maximizing power of relevance tests.

## Data Availability

The simulated data and the scripts that support the evaluation of the proposed method are openly available online at https://doi.org/10.5281/zenodo.13692989.

## Acknowledgments

This work was financed by a CIFRE agreement (Conventions Industrielles de Formation par la Recherche) of the ANRT (Association Nationale de la Recherche et de la Technologie). The CIFRE agreement is a partnership between a public laboratory and a company, here the UMR (Unité Mixte de Recherche) 1137 and Ipsen, respectively. The authors would like to thank the entire PFIM group, and in particular Romain Leroux, PFIM6 development manager of PFIM6, for their help in adapting the R code for our use.

## Conflict of interest

L.F and K.B. are full-/-part-time employees of Ipsen.

## A Technical appendix

### A.1 Power of Test

#### A.1.1 Power computation for significance test

The null hypothesis for the Wald test of comparison is *H*_0_ : *β* = 0 and the alternative hypothesis is *H*_1_ : *β* ≠ 0. The test statistics is given by equation (23).

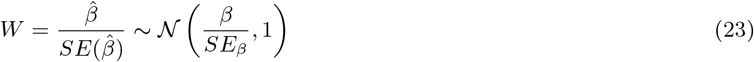

- Under *H*_0_, *W* ∼ 𝒩 (0, 1), therefore *H*_0_ is rejected if ❘*W❘ q*_1−*α/*2_, where *q*_1 − *α/*2_ denotes the 1 − *α/*2-quantile of the normal distribution
- Under *H*_1_, 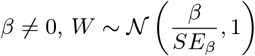

The power is computed for a given *β* as the probability to reject *H*_0_ under *β ∈H*_1_:

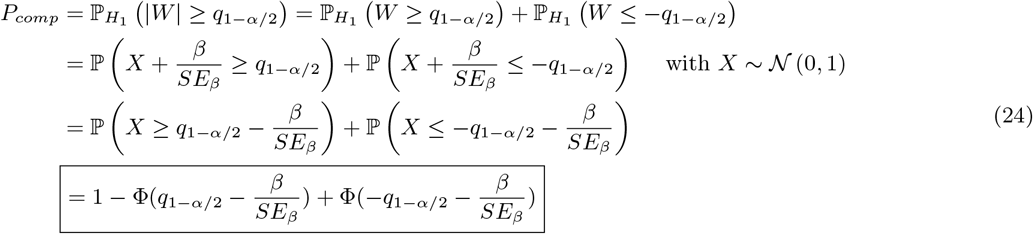

- *β* > 0: Because 𝕡 *X* ≤ −*q*_1−*α/*2_ and 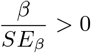, we have that 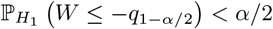 Therefore, the second term can be neglected and

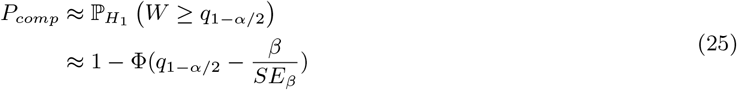
- *β* < 0: On the contrary,

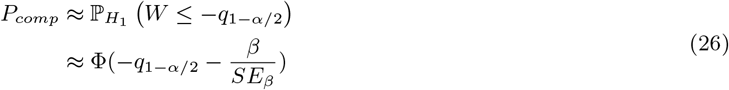

#### A.1.2 Power computation for Relevance test

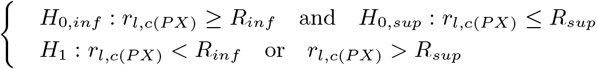

Thus *H*_0_ is not rejected unless neither *H*_0,*inf*_ nor *H*_0,*sup*_ is rejected.

For 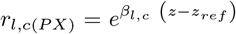, the hypothesis write: *H*_0_ : *β*_*l,c*_ *∈* [*B*_*inf*_ ; *B*_*sup*_], where depending on the sign of (*z* − *z*_*ref*_), *B*_*sup*_ and *B*_*inf*_ equals 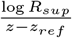 or 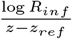.

The null hypothesis is rejected if *H*_0,*inf*_ is rejected, i.e.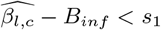, or if *H*_0,*sup*_ is rejected, i.e.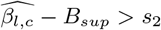 ; where *s*_1_ and *s*_2_ are chosen to reach an *α* type one error. Because these two events are incompatible the following holds:

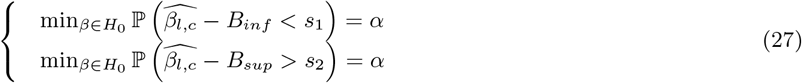

Recalling that 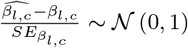, the system writes:

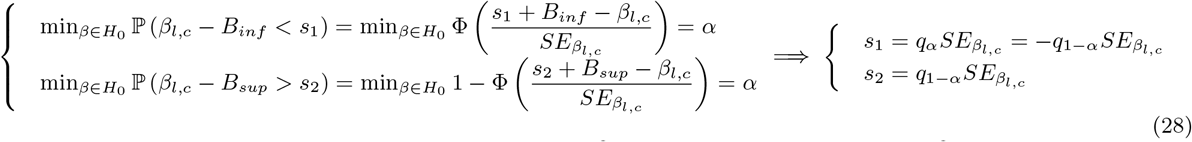

Finally, at level 1 − *α*, the null hypothesis is rejected if 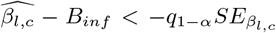 or if 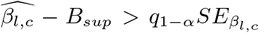 ; equivalently, the null hypothesis is rejected if 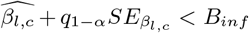 or if 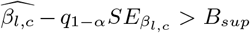. The first inequality involves the upper bound of the 1 − 2*α* CI on the ratio, while the second inequality involves the lower bound.

The power is the probability under *H*_1_ to reject *H*_0_:

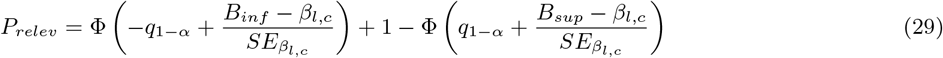

On one hand, if 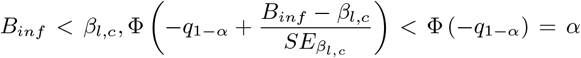, therefore this term is negligible in power computation. On the other hand, if 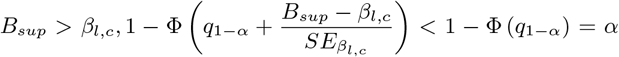, therefore this term is negligible in power computation. Consequently, to compute the number of subjects needed to achieve, a given power let us distinguish two cases:

- If 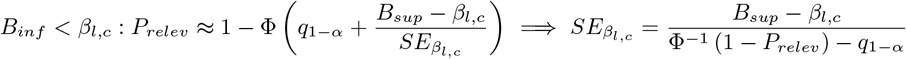
- If 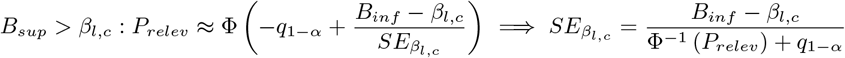

## B Addition details on the evaluation by simulation

**Figure 4:**
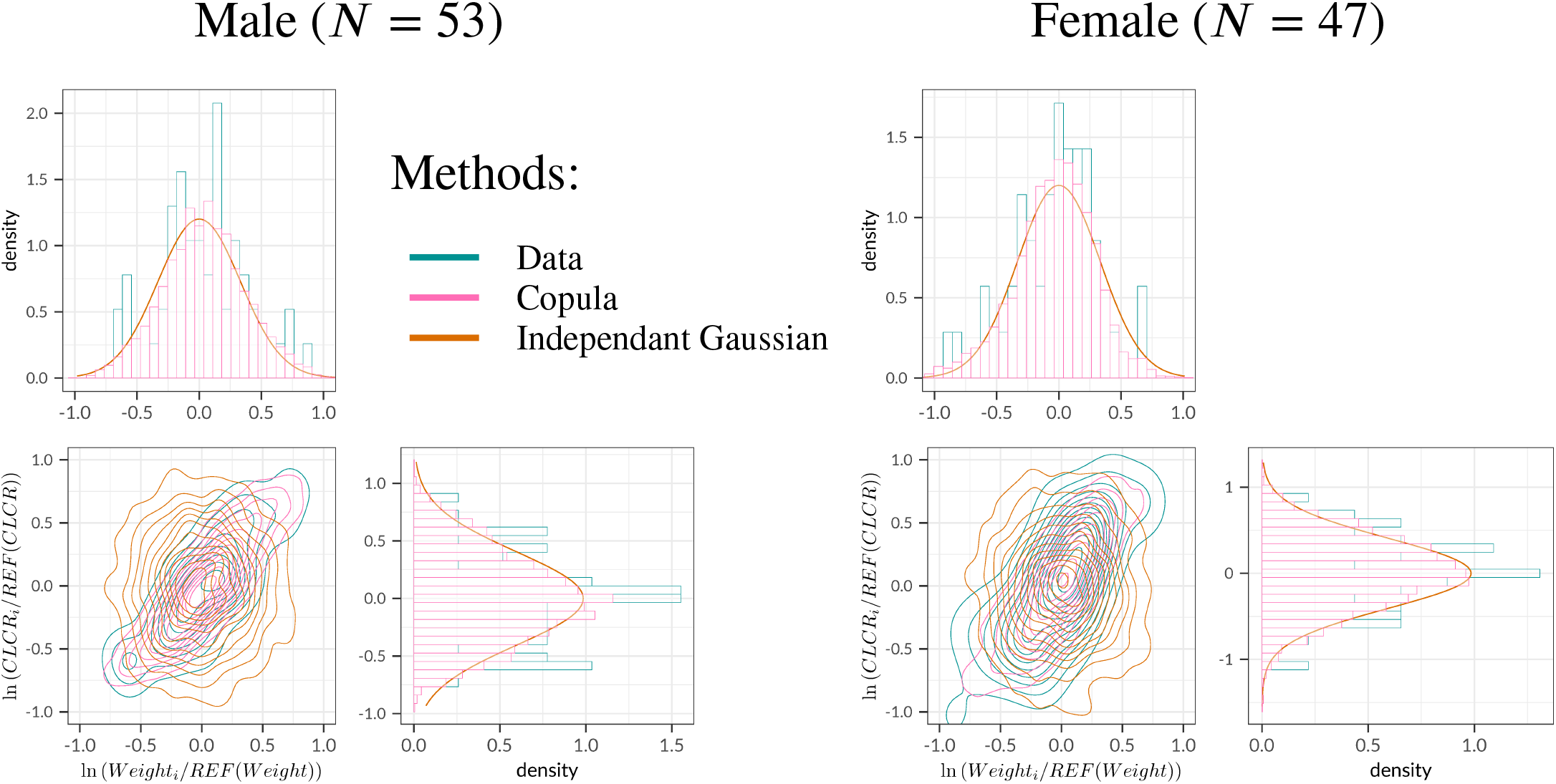
Evaluation - Covariate distributions for Male on the left and Female on the right: Data boxplots, fitted Copulas boxplots and fitted independent Gaussian densities.

**Figure 5:**
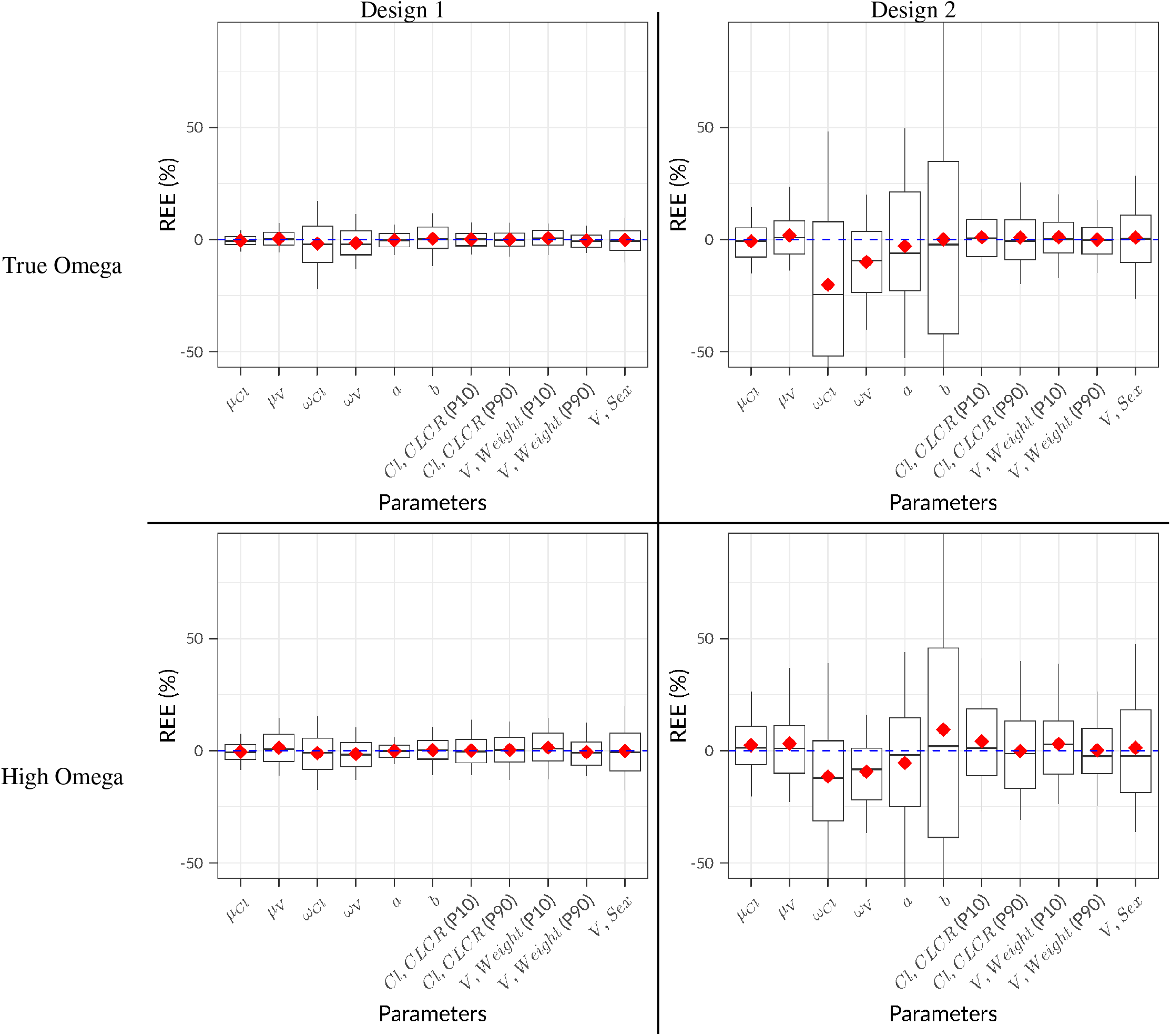
Evaluation - Boxplots of Relative Estimation Error across 200 datasets for the 4 scenarios The boxplot displays the median, the 25th and 75th percentiles, while the whiskers are 5th and 95th percentiles. The red diamond corresponds to the mean.

**Figure 6:**
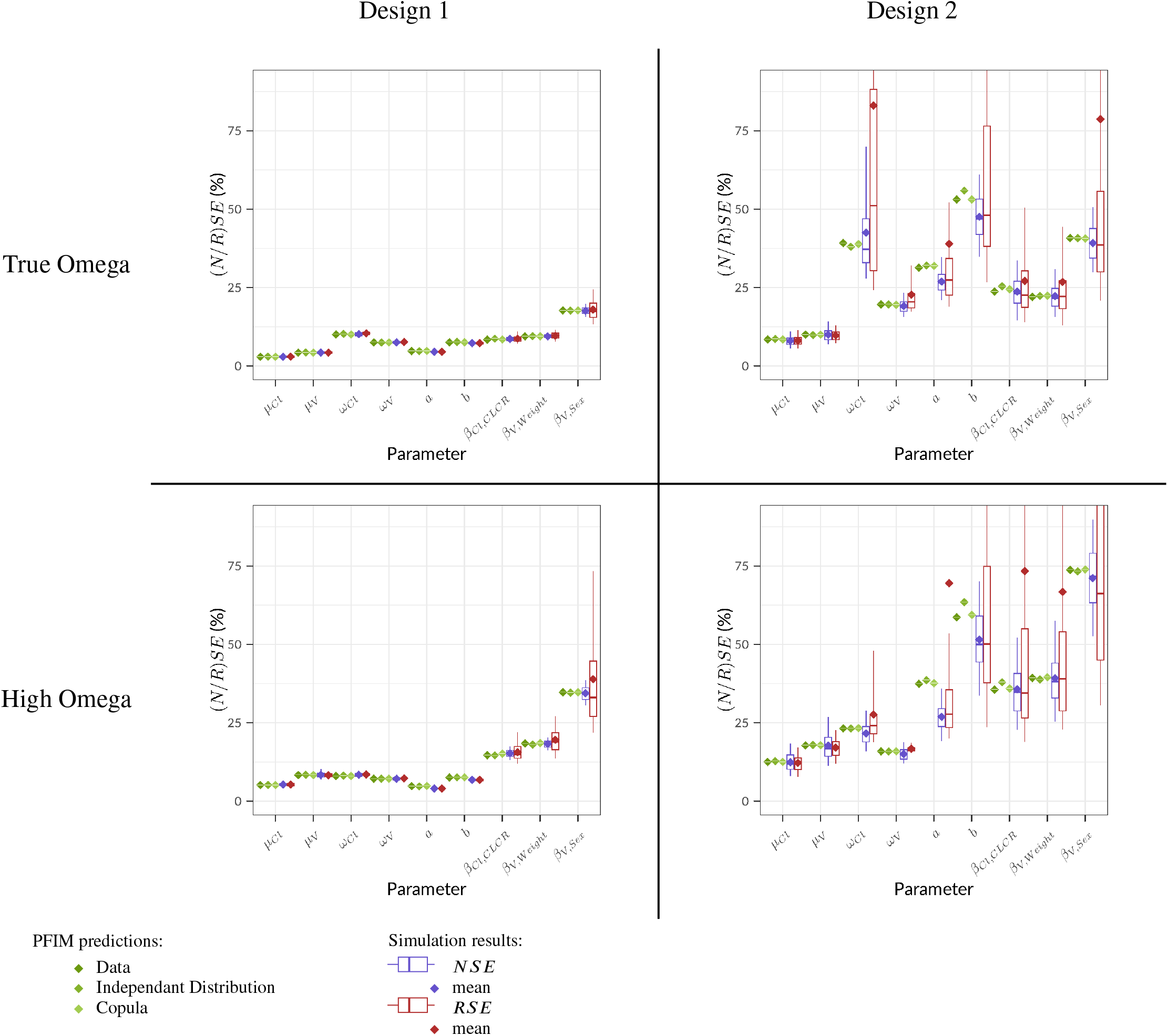
Evaluation - Normalized and Relative Standard Error for the 4 scenarios: PFIM predictions using the three methods for handling covariates and simulation results across 200 datasets The boxplot displays the median, the 25th and 75th percentiles, while the whiskers are 5th and 95th percentiles.

## C Addition details on PK example simulation

**Table 8:** Application - Theoretical Sampling Design.

| Study | $N$ | Daily Doses, mg ($N$) | $n$ | Samplings, hours after first dose |
| --- | --- | --- | --- | --- |
| XL184-001 | 40 | 140 (35) - 200 (5) | 15 | 0.5, 1, 2, 4, 8, 96, 100, 336, 432, 432.5, 433, 434, 436, 440, 672 |
| XL184-010 | 77 | 140 (77) | 43 | 0.5, 1, 2, 3, 4, 5, 6, 8, 10, 12, 14, 24, 48, 72, 120, 168, 240, 288, 336, 408, 504, 744, 744.5, 745, 746, 747, 748, 749, 750, 752, 754, 756, 758, 768, 792, 816, 864, 912, 984, 1032, 1080, 1152, 1248 |
| XL184-020 | 63 | 20 (21) - 40 (21) - 60 (21) | 21 | 0.5, 1, 2, 3, 4, 5, 6, 8, 10, 12, 14, 24, 48, 72, 120, 168, 240, 288, 336, 408, 504 |
| XL184-201 | 39 | 140 (39) | 7 | 336, 340, 672, 676, 1008, 1012, 1344 |
| XL184-203 - NRE | 136 | 40 (55) - 60 (36) - 100 (45) | 5 | 504, 1008, 2016, 3024, 4032 |
| XL184-203 - RDT | 185 | 60 (35) - 100 (150) | 5 | 2016, 2352, 2688, 3024, 3360 |
| XL184-301 | 210 | 100 (1) - 140 (209) | 7 | 2, 4, 6, 672, 674, 676, 678 |
| XL184-306 | 41 | 20 (1) - 40 (1) - 60 (39) | 3 | 504, 1008, 2016 |
| XL184-307 | 498 | 20 (18) - 40 (41) - 60 (439) | 2 | 504, 2016 |
| XL184-308 | 282 | 20 (5) - 40 (33) - 60 (244) | 2 | 672, 1344 |
| XL184-309 | 452 | 40 (7) - 60 (445) | 3 | 336, 672, 1344 |

$N$ : refers to the number of subjects included in the study, $n$ : the number of PK samplings per subject

**Figure 7:**
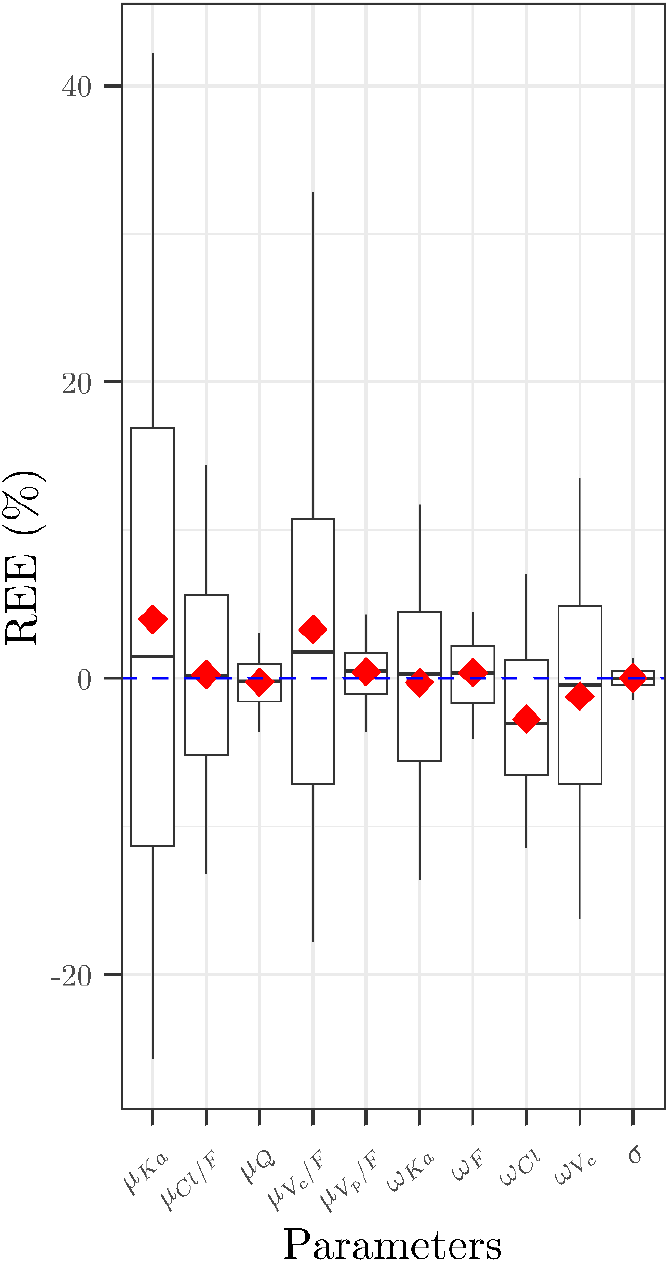
Application - Boxplots of Relative Estimation Error across 200 datasets - Base parameters The boxplot displays the median, the 25th and 75th percentiles, while the whiskers are 5th and 95th percentiles. The red diamond corresponds to the mean.

**Figure 8:**
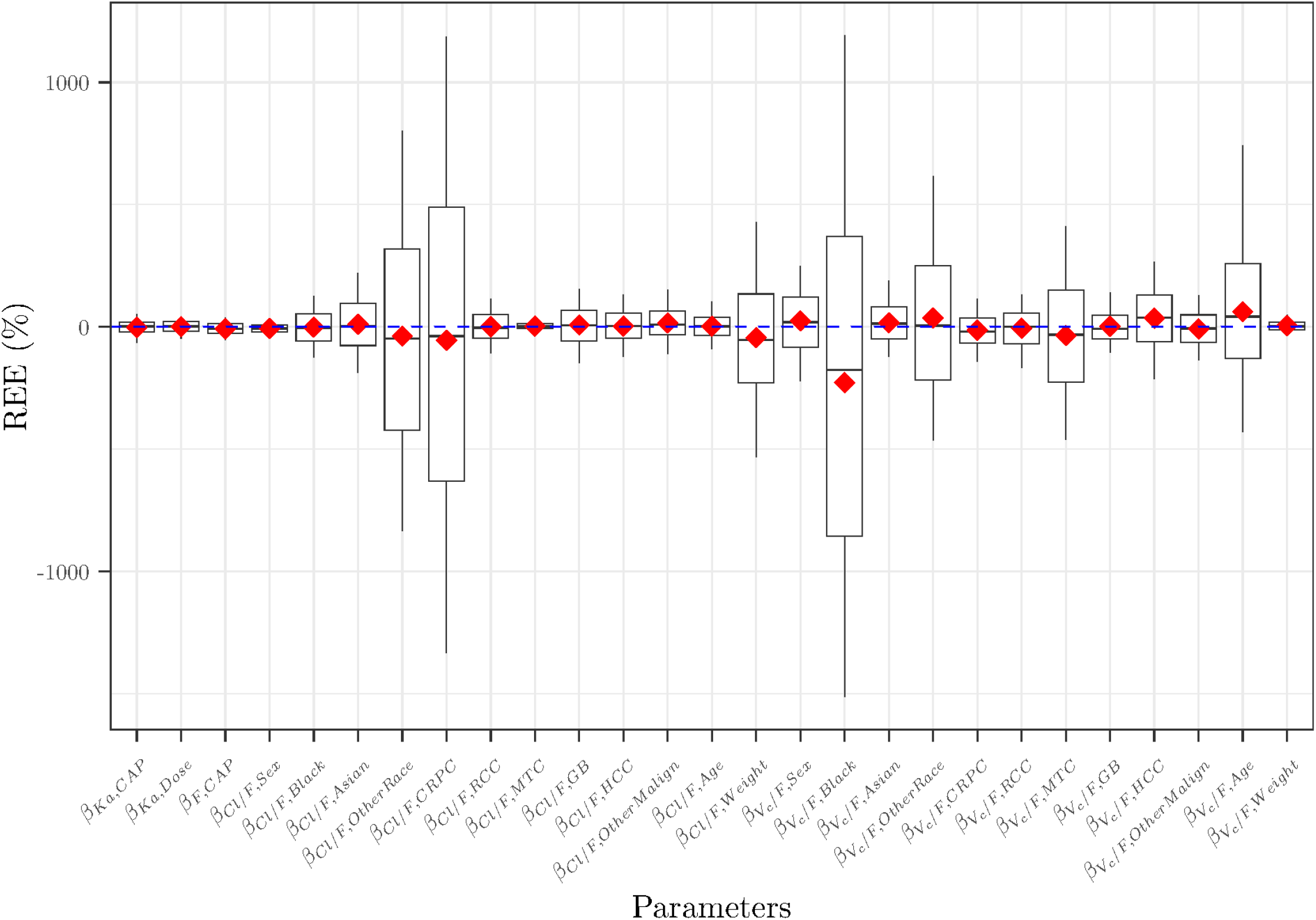
Application - Boxplots of Relative Estimation Error across 200 datasets - Covariate effects parameters The boxplot displays the median, the 25th and 75th percentiles, while the whiskers are 5th and 95th percentiles. The red diamond corresponds to the mean.

**Table 9:** Application - *RB* and *RRMSE* across 200 datasets - Base parameters.

| Parameter | RB (%) | RRMSE (%) |
| --- | --- | --- |
| $\mu_{Ka}$ | 3.97 | 20.9 |
| $\mu_{Cl/F}$ | 0.237 | 7.90 |
| $\mu_Q$ | -0.282 | 2.02 |
| $\mu_{V_c/F}$ | 3.27 | 16.1 |
| $\mu_{V_p/F}$ | 0.418 | 2.37 |
| $\omega_{Ka}$ | -0.305 | 7.85 |
| $\omega_F$ | 0.36 | 2.58 |
| $\omega_{Cl}$ | -2.77 | 6.47 |
| $\omega_{V_c}$ | -1.24 | 9.15 |
| $\sigma$ | 0.00625 | 0.797 |

**Table 10:** Application - Simulation Value, Estimate, *RB* and *RRMSE* across 200 datasets - Covariate effects parameters.

| Parameter | Value | Estimate | RB (%) | RRMSE (%) |
| --- | --- | --- | --- | --- |
| $\beta_{Ka,CAP}$ | -0.911 | -0.892 | -2.11 | 33.9 |
| $\beta_{Ka,Dose}$ | 0.734 | 0.738 | 0.578 | 29.1 |
| $\beta_{F,CAP}$ | -0.166 | -0.153 | -7.74 | 30.8 |
| $\beta_{Cl/F,Sex}$ | -0.274 | -0.257 | -6.17 | 23.6 |
| $\beta_{Cl/F,Black}$ | 0.162 | 0.16 | -1.08 | 83.0 |
| $\beta_{Cl/F,Asian}$ | -0.0668 | -0.0741 | 10.9 | 127 |
| $\beta_{Cl/F,OtherRace}$ | 0.0279 | 0.0174 | -37.8 | 537 |
| $\beta_{Cl/F,CRPC}$ | -0.0115 | -0.00522 | -54.6 | 808 |
| $\beta_{Cl/F,RCC}$ | -0.139 | -0.14 | 0.361 | 67.0 |
| $\beta_{Cl/F,MTC}$ | 0.643 | 0.661 | 2.80 | 16.2 |
| $\beta_{Cl/F,GB}$ | 0.178 | 0.192 | 7.73 | 96.7 |
| $\beta_{Cl/F,HCC}$ | -0.13 | -0.134 | 3.38 | 76.0 |
| $\beta_{Cl/F,OtherMalign}$ | 0.171 | 0.199 | 16.3 | 83.4 |
| $\beta_{Cl/F,Age}$ | -0.157 | -0.161 | 2.41 | 61.0 |
| $\beta_{Cl/F,Weight}$ | -0.0393 | -0.0221 | -43.8 | 285 |
| $\beta_{V_c/F,Sex}$ | 0.0939 | 0.117 | 24.4 | 148 |
| $\beta_{V_c/F,Black}$ | 0.0441 | -0.0564 | -228 | 930 |
| $\beta_{V_c/F,Asian}$ | -0.363 | -0.423 | 16.5 | 95.4 |
| $\beta_{V_c/F,OtherRace}$ | -0.126 | -0.172 | 36.5 | 362 |
| $\beta_{V_c/F,CRPC}$ | -0.297 | -0.256 | -13.9 | 82.2 |
| $\beta_{V_c/F,RCC}$ | -0.422 | -0.397 | -5.88 | 91.4 |
| $\beta_{V_c/F,MTC}$ | -0.0657 | -0.0425 | -35.3 | 278 |
| $\beta_{V_c/F,GB}$ | -0.735 | -0.75 | 2.09 | 76.0 |
| $\beta_{V_c/F,HCC}$ | -0.166 | -0.225 | 35.7 | 158 |
| $\beta_{V_c/F,OtherMalign}$ | -0.272 | -0.248 | -8.8 | 85.5 |
| $\beta_{V_c/F,Age}$ | 0.0644 | 0.104 | 62.1 | 347 |
| $\beta_{V_c/F,Weight}$ | 1.19 | 1.25 | 4.74 | 25.6 |

CRPC: Castration-resistant prostate cancer, RCC: Renal cell carcinoma, MTC: Metastatic medullary thyroid cancer, GB: Glioblastoma multiforme, HCC: Hepatocellular carcinoma, OtherMalign: other malignancy, CAP: Capsule formulation

**Figure 9:**
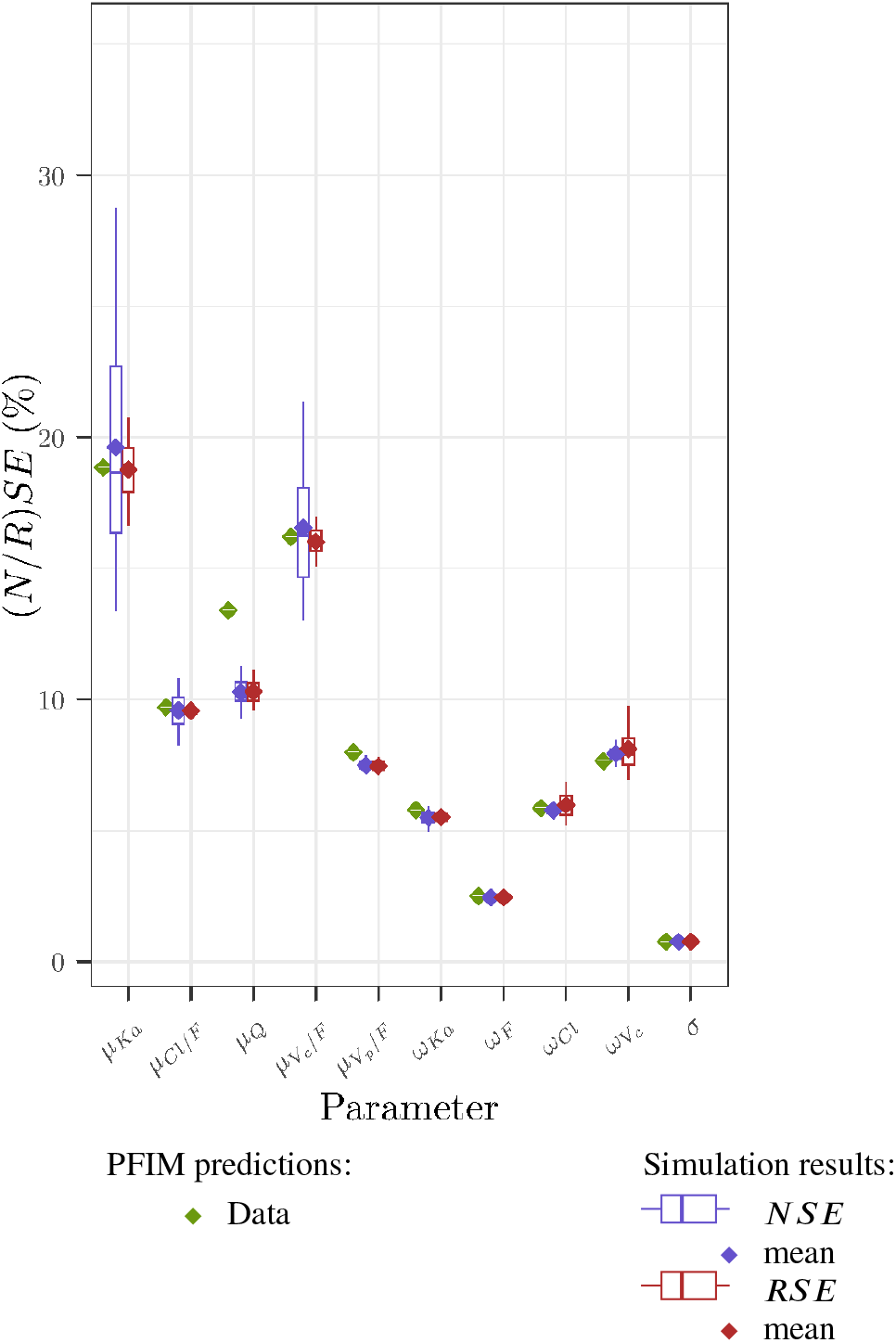
Application - Normalized and Relative Standard Error - Base parameters: PFIM predictions using the Data method for handling covariates and simulation results across 200 datasets The boxplot displays the median, the 25th and 75th percentiles, while the whiskers are 5th and 95th percentiles.

**Figure 10:**
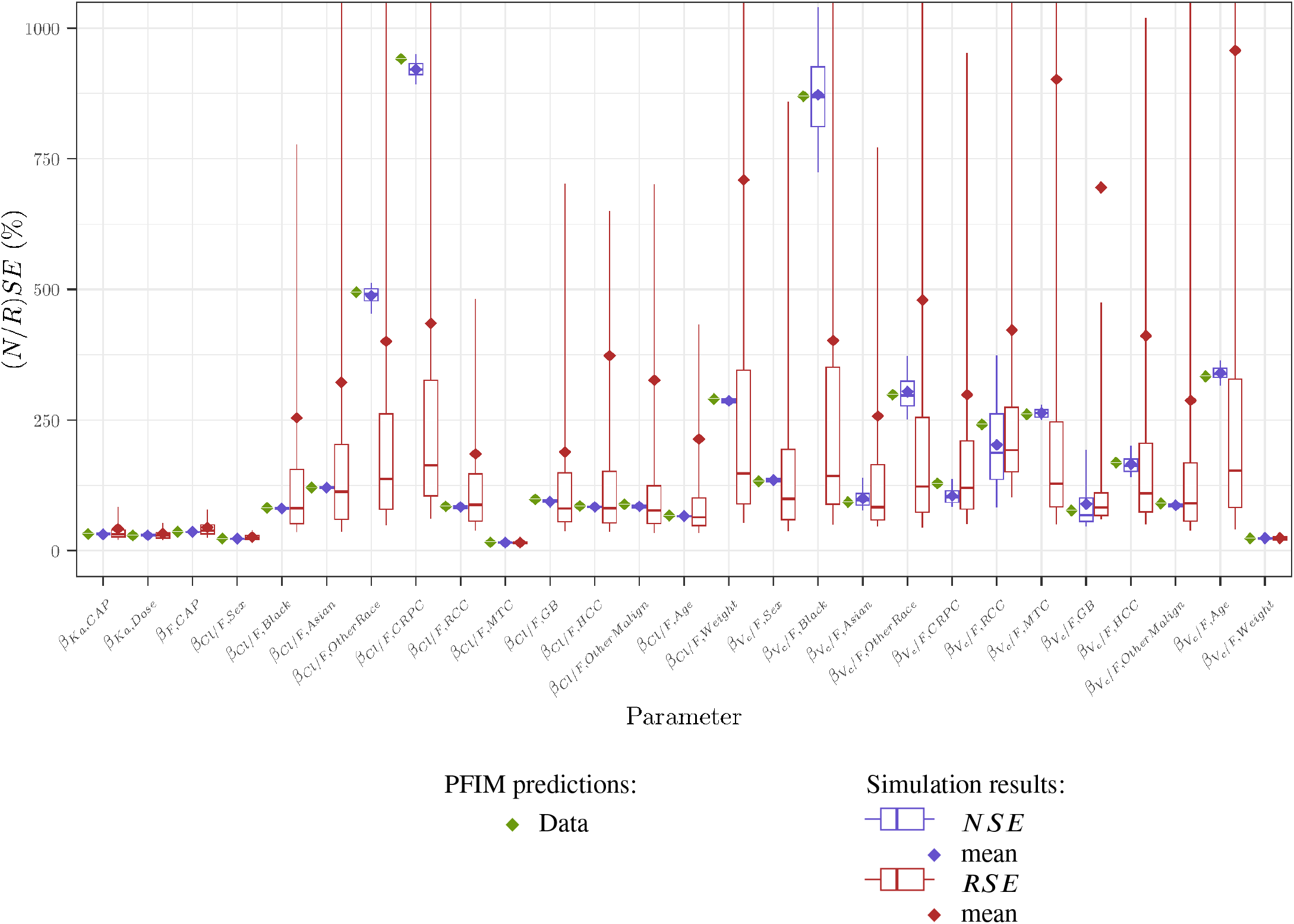
Application - Normalized and Relative Standard Error - Covariate effects: PFIM predictions using the Data method for handling covariates and simulation results across 200 datasets The boxplot displays the median, the 25th and 75th percentiles, while the whiskers are 5th and 95th percentiles.

## Notes

### Author Declarations

This work used the same data as "Nguyen, L., Chapel, S., Tran, B. D., & Lacy, S. (2019). Updated population pharmacokinetic model of cabozantinib integrating various cancer types including hepatocellular carcinoma. The Journal of Clinical Pharmacology, 59(11), 1551-1561." The latter pooled data from 10 clinical trials for which all protocols were approved by institutional review boards of participating institutions, and written informed consent was obtained from all HV and patients prior to enrollment.

